# Retest-Reliability of Cone and Rod Function Assessments in Pseudoxanthoma elasticum: PROPXE Study Report 3

**DOI:** 10.64898/2025.12.15.25342250

**Authors:** Giuseppe Cancian, Georg Ansari, Chantal Dysli, Stephan Michels, Nicolas Feltgen, Sharon F. Terry, Maximilian Pfau, Kristina Pfau, the PROPXE Study Group

## Abstract

**Purpose:** To determine the test-retest reliability of visual function parameters in patients with genetically confirmed Pseudoxanthoma elasticum (PXE), as a necessary step toward evaluating their suitability as outcome measures in future therapeutic trials.

**Methods:** In this prospective natural history study (PROPXE, ClinicalTrials.gov ID: NCT05662085), patients with PXE underwent comprehensive visual function evaluation in one study eye at baseline and at a month 2 retest visit. Functional testing included light- and dark-adapted steady state microperimetry and dark-adaptometry at 8°, 15° 30°, and 46° eccentricity. Test-retest reliability was evaluated using Bland-Altman statistics.

**Results:** Twenty-six patients (14 female, 12 male; median [IQR] age 55 years [43; 59]) with genetically confirmed PXE were included in the study. Overall, the steady-state microperimetry limits of agreement (LoA) were ±2 dB for mean sensitivity and ±6.8 dB for pointwise sensitivity in both scotopic (cyan and red) and mesopic conditions. The LoAs of rod intercept time as a measure of dark adaptometry were ± 12 min at the inner measurement points (8° and 15°) and ± 18 min at the outer measurement points (30° and 46°).

**Conclusions:** Scotopic and mesopic microperimetry LoAs are similar to earlier published test-retest analyses in other retinal diseases. Dark-adaptometry curve parameters were markedly more variable compared to previous data in healthy volunteers. This is likely attributable to the severe dark adaptation abnormalities in PXE, leading to long test durations.

**Translational Relevance:** The evaluation of functional biomarkers is critical for designing future clinical trials aimed at slowing PXE progression.

## INTRODUCTION

Due to advancements in technology and the widespread availability of genetic testing, along with the emerging potential of new gene therapies, the evaluation of patients with degenerative and inherited retinal diseases has undergone a significant transformation over the past decade. In a subset of (predominantly) macular diseases with early alterations at the level of Bruch’s membrane (BrM), localized psychophysical measures of dynamic cone and/or rod dark adaptation may reveal early dysfunction, while the steady-state function is still intact.^1–3^ This includes common diseases such as age-related macular degeneration, as well as inherited retinal diseases (IRDs) like Pseudoxanthoma elasticum (PXE), Sorsby fundus dystrophy, and late-onset retinal degeneration.^3–5^

PXE is a disease with primary alterations at the level of BrM. It is an autosomal recessive disease caused by mutations in the *ABCC6* gene, leading to calcifications of elastic and collagen fibers in connective tissues throughout the body.^6–8^ Therefore, several organ systems are affected, primarily the eyes, the skin, and the vascular system. In the eye, PXE causes BrM calcification that progresses centrifugally over time.^8–10^ The calcified BrM is already present in the initial stage of the disease as an area with ’granular’ or ’dotted’ aspect at the posterior pole of the eye (’Peau d’orange’). Eventually, with centrifugal progression of calcification, the central part of Peau d’orange coalesces, forming a central area of continuously calcified BrM (’Coquille d’œuf’) (Figure 1).^11^ With disease progression, the calcification of BrM leads to secondary complications including angioid streaks, macular neovascularization (MNV), and atrophy of the outer retina and retinal pigment epithelium (RPE).

**Figure 1.**
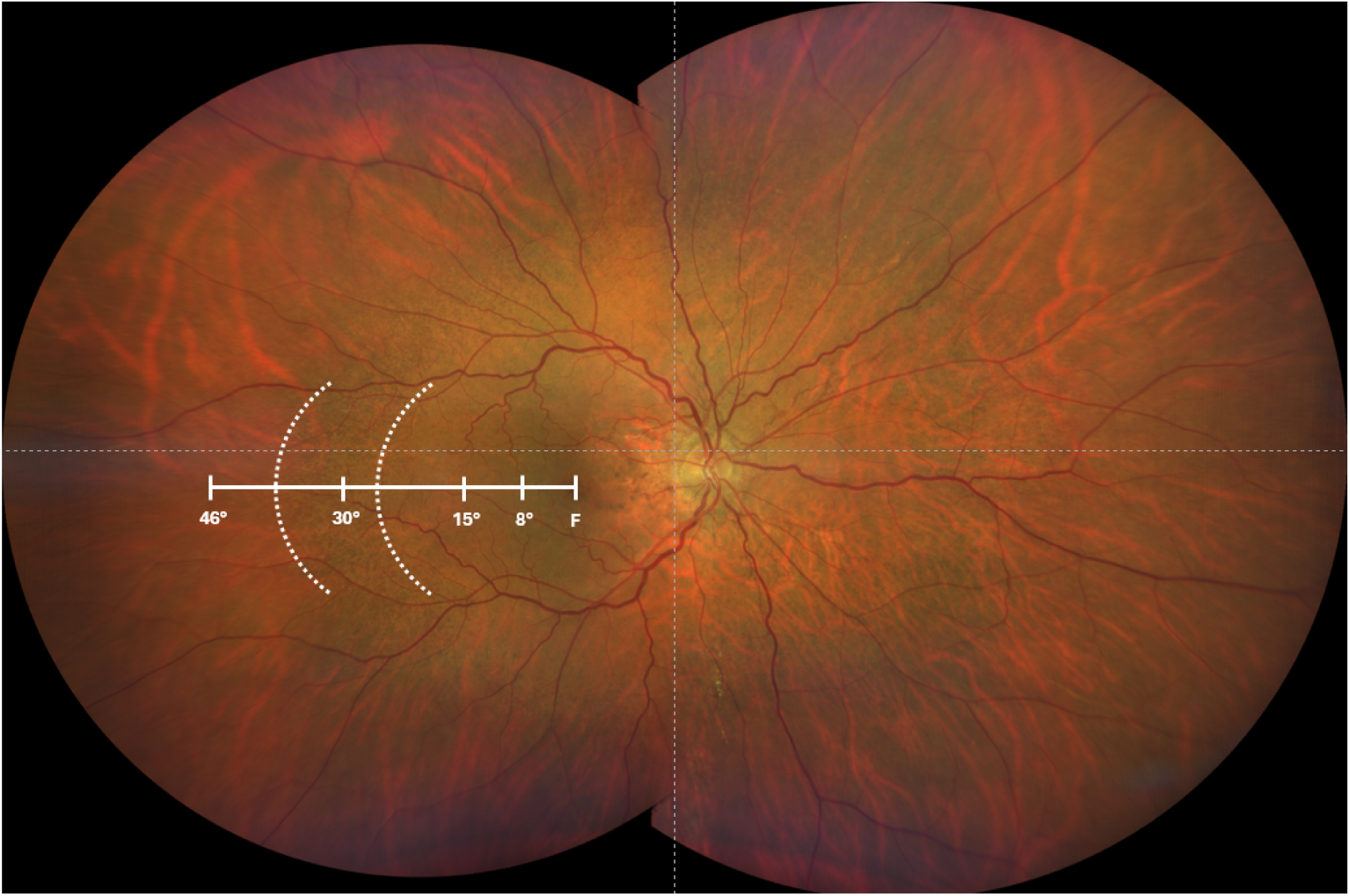
Fundus photography of the retina from a patient with PXE. The dashed white lines delineate the temporal outer and inner boundaries of the Peau d’orange, representing the transition zone between the peripheral noncalcified Bruch’s membrane and the central continuously calcified Bruch’s membrane. The solid white lines indicate the fovea (F) and the retinal eccentricities where dark adaptometry was performed: at 8° and 15°, within the continuously calcified region; at 30°, inside the Peau d’orange; and at 46°, outside the Peau d’orange.

Recent evidence has shown that a delayed rod-mediated dark adaptation is one of the earliest indicators of functional impairment and may serve as a valuable metric for evaluating interventions aimed at slowing BrM calcification.^12^ Despite this potential and recent as well as imminent treatment trials (e.g., clinicaltrials.gov NCT04868578, NCT05832580), there is a notable lack of retest reliability data for dark adaptometry parameters, especially PXE-relevant retinal loci outside of the macula. Steady-state loss of rod function as measured by scotopic microperimetry is likely also a relatively early form of dysfunction preceding the loss of cone function.^5^ However, retest-reliability in a disease-specific context is also lacking. This gap in the literature underscores the need for further research to establish the consistency and reliability of rod-mediated dark adaptation delays and other visual function tests as potential biomarkers in these diseases.

Thus, the objective of the present study was to perform a comprehensive evaluation of test-retest reliability across all visual function metrics assessed within the framework of the PROPXE study. Specifically, we quantified the reproducibility of best-corrected visual acuity (BCVA), contrast sensitivity function testing, steady-state light- and dark-adapted microperimetry, and parameters derived from dynamic dark adaptation testing. In addition, we performed a literature review and comparison against previously published reliability data for analogous measures in other inherited and acquired retinal diseases, including light- and dark-adapted perimetry, microperimetry, and dark adaptometry (see Supplementary Table S1).

## METHODS

This study included individuals from the prospective natural history study ’Progression Rate of Pseudoxanthoma Elasticum-associated Choroidal and Retinal Degeneration’ (PROPXE, ClinicalTrials.gov ID: NCT05662085).^13^ The study was approved by the authorized human research ethics committee (EKNZ) and adhered to the tenets of the Declaration of Helsinki. All participants were informed of the study’s nature and provided written informed consent before participating in study-related examinations.

### Study Design and Core Examinations

The study design, as well as the inclusion and exclusion criteria, have been described in detail previously.^13^ This study involves a baseline visit and a retest, with follow-up examinations planned for year one and year two. It is currently ongoing.

Participants underwent comprehensive ophthalmic evaluations, including BCVA assessments using the qVA protocol on the Manifold platform (Adaptive Sensory Technology, Lübeck, Germany), as well as quick contrast sensitivity function testing (qCSF) on the same platform.

A panel of standardized multimodal imaging was performed. Spectral-domain optical coherence tomography (SD-OCT) imaging of the macula was obtained using a Heidelberg Spectralis device (Heidelberg Engineering, Heidelberg, Germany) with a 30° x 25° field of view (121 B-scans, HR mode, enhanced Automatic Real-Time Function [ART] 25). In addition, 55° fundus autofluorescence (FAF) and 9-gaze infrared reflectance (IR) images were obtained. Color fundus photography (CFP) was obtained using a Clarus 700 imaging device (Carl Zeiss Meditec AG, Jena, Germany) with the ultra-widefield mode.

### Light- and Dark-Adapted Visual Function Assessments

For visual function assessments, one study eye was selected for each patient. The treatment-naïve eye (i.e., eyes with no history of exudative macular neovascularization [MNV]) was preferred. If both or no eyes had a history of exudative MNV, the eye with better acuity was chosen. If acuity was also identical, the right eye was chosen. Retinal sensitivity of the posterior pole was examined using the fundus-controlled perimetry (microperimetry)^14^ device S-MAIA (CenterVue/iCare, Padua, Italy). First, light-adapted mesopic microperimetry was performed using a 4-2 projection strategy and a pattern of 61 Goldmann III-sized stimuli along the horizontal meridian through the fovea, covering 15° to the temporal and 15° to the nasal side (Supplementary Figure S1).

Dark adaptation was performed for 45 minutes. After dark adaptation, a dark-adapted two-color microperimetry (S-MAIA) test was performed using the same strategy and grid as described above.

Dark adaptometry testing (MonCvONE)^15^ was conducted after an initial bleaching protocol involving a full-field 634 photopic cd/m^2^ (946 scotopic cd/m^2^) bleach for 5 minutes, corresponding to a 59% rhodopsin bleach. Dark adaptometry testing was performed with cyan and red Goldmann V-sized stimuli (peak wavelengths: 500 nm and 647 nm, stimulus duration: 200 ms) at 8°, 15°, 30°, and 46° eccentricity temporal to the fixation locus (equivalent to the nasal visual field). The temporal retina was selected to avoid the optic nerve head and the parapapillary region (i.e., the region where MNV-related atrophy often first manifests in PXE). The four test loci were designated to measure within the continuously calcified BrM (8°), in proximity to the Peau d’orange inner boundary (15°) and outer boundary (30°), and outside of the calcified BrM (46°). Dark adaptometry testing was conducted for up to 60 minutes with the option to terminate the test early if all four loci reached their final steady-state threshold before 60 minutes.

## Statistical Analyses

All statistical analyses were performed in *R* using the add-on packages *tidyverse*,^16^ ggplot2^17^ and *dplyr*.^18^ Test-retest reliability was calculated using the *R* package *SimplyAgree*.^19,20^

qCSF acuity provided in cycles per degree (cpd) was converted to logMAR using the following formula:

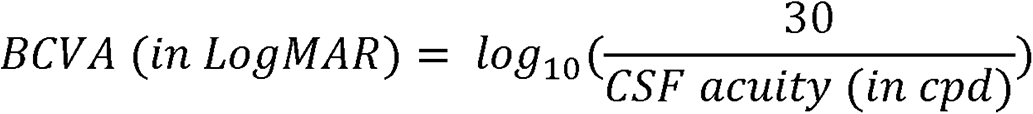

Test–retest reliability of mean sensitivity between baseline and retest visits was assessed using Bland–Altman analysis. The 95% confidence interval (CI) of the mean bias — defined as the average difference between paired measurements (month 2 ‘retest’ - baseline) — was calculated using the formula:

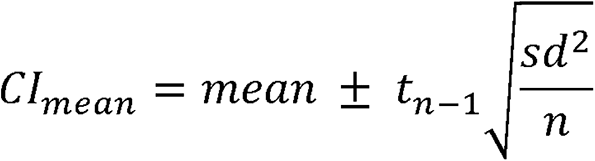

where *t* denotes the Student’s t distribution and *n* the cardinality of the sample.

The limits of agreement (LoA) were determined as the mean bias ± 1.96 times the standard deviation (SD) of the differences. The corresponding 95% CIs for the upper and lower LoA were derived using:

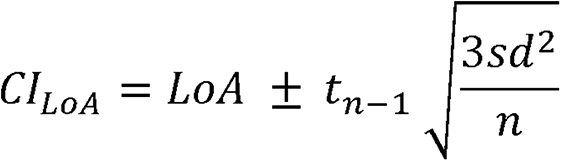

Bland–Altman plots illustrate the mean bias (with 95% CI) along with the upper and lower LoA (also plotted with outer 95% CI). For pointwise microperimetry analysis, stimulus values were nested within patient IDs to avoid spuriously narrow LoA estimates due to within-subject clustering.

Last, we complied comparable test-retest reliability data for dark adaptometry data and microperimetry across diseases (Supplementary Table S1)^15,21–35^.

## RESULTS

### Patient Characteristics

A total of 52 eyes of 26 patients (14 females, 54%) with a median (IQR) age of 55 (43; 59) years were included in the study (Table 1). Of the 26 study eyes, 14 study eyes (54%) had a history of exudative MNV.

**Table 1.**
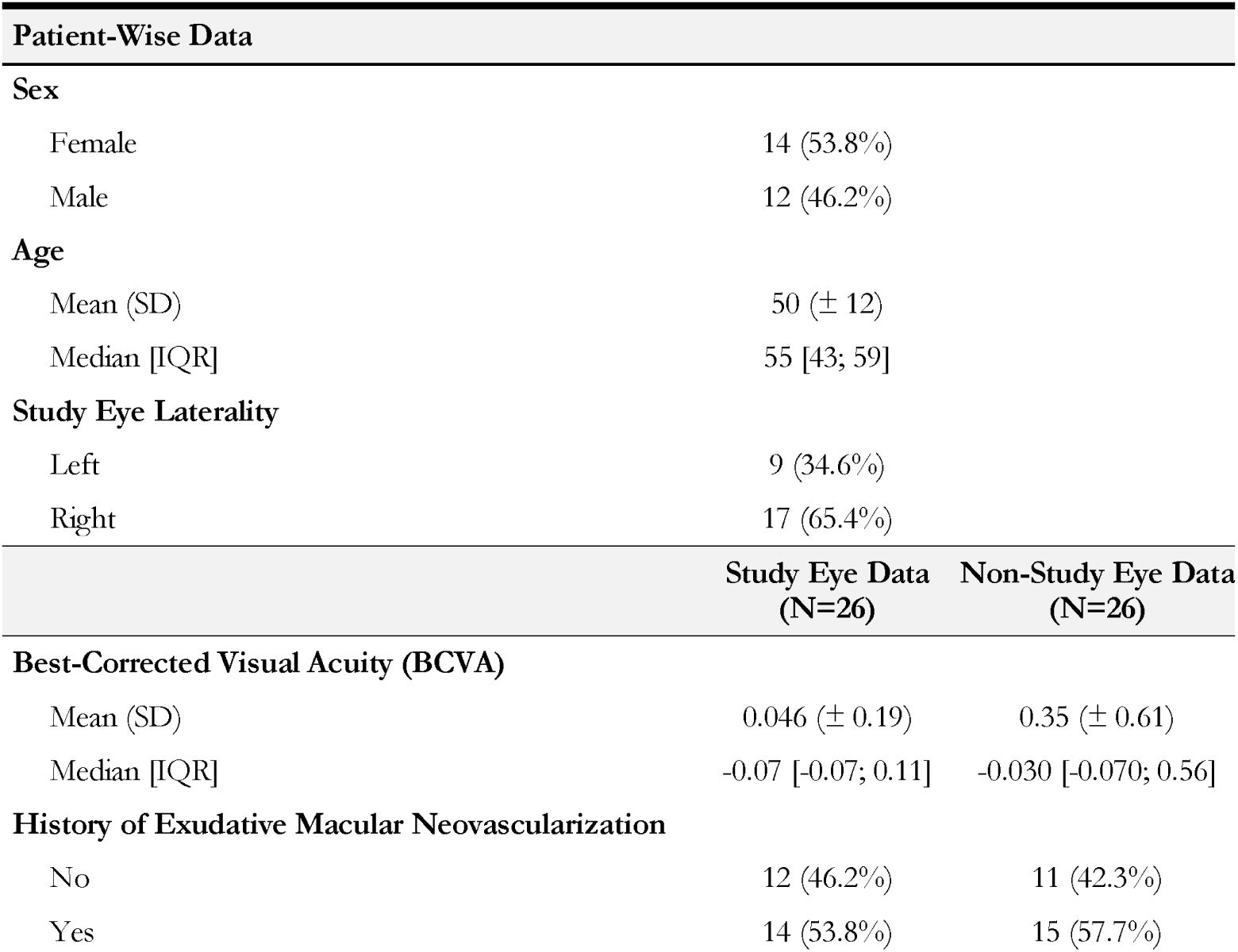
Demographics

The median interval between the baseline and retest visit was 58 [IQR: 7.75] days.

### Functional Disease Stability

Between the baseline exam and the retest exam, there were no significant changes in median visual acuity, AULCSF, or CSF Acuity between the baseline and second visits for either eye (all p-values > 0.05; Table 2). These findings indicate that basic visual functions remained stable during the study period. Therefore, any variations observed in the dark adaptometry and fundus-tracked microperimetry tests are likely due to test-retest variability rather than actual clinical changes.

**Table 2.** Chart-based vision tests results

|  | Baseline<sup>a</sup> | Second visit<sup>a</sup> | Wilcoxon signed-rank test |
| --- | --- | --- | --- |
| Visual Acuity (RE, logMAR) | -0.01 [-0.07; 0.35] | -0.03 [-0.07; 0.25] | $P = 0.610$ |
| Visual Acuity (LE, logMAR) | -0.07 [-0.07; 0.11] | -0.07 [-0.07; 0.11] | $P = 0.265$ |
| AULCSF (RE, logCS*logCPD) | 1.25 [0.35; 1.36] | 1.17 [0.63; 1.39] | $P = 0.833$ |
| AULCSF (LE, logCS*logCPD) | 1.34 [1.09; 1.44] | 1.33 [1.08; 1.44] | $P = 0.191$ |
| CSF Acuity (RE, logMAR) | 0.20 [0.09; 0.42] | 0.17 [0.09; 0.32] | $P = 0.211$ |
| CSF Acuity (LE, logMAR) | 0.13 [0.05; 0.28] | 0.14 [0.05; 0.31] | $P = 0.900$ |

<sup>a</sup>: values are expressed as median [interquartile range (IQR)]
RE: right eye; LE: left eye; AUL: area under the log; CSF: contrast sensitivity function; logMAR: logarithm of the minimum angle of resolution.

### Retest-Reliability of Chart-Based Vision Tests

For qVA-based best-corrected visual acuity, the CoR was 0.303 logMAR for the right eye (OD) and 0.146 logMAR for the left eye (OS). The LoA ranged from −0.306 logMAR to 0.300 logMAR for OD and from −0.171 logMAR to 0.121 logMAR for OS, indicating good repeatability between the two visits.

For qCSF Acuity, the CoR was 0.265 logMAR (OD) and 0.191 logMAR (OS). The LoA ranged from -0.288 logMAR to 0.241 logMAR for OD and from -0.200 logMAR to 0.182 logMAR for OS, indicating good repeatability in test-retest measurements also for this parameter.

For the qCSF-based AULCSF, the CoR was 0.323 logCS*logCPD (OD) and 0.247 logCS*logCPD (OS). The LoA extended from −0.323 to 0.322 logCS*logCPD for OD and from −0.219 to 0.274 logCS*logCPD for OS, suggesting acceptable repeatability for these measurements.

The detailed results of the test-retest variability parameters are listed in Table 3. The Bland-Altman plots for the six variables of the Chart-Based Vision Tests are shown in Figure 2.

**Figure 2.**
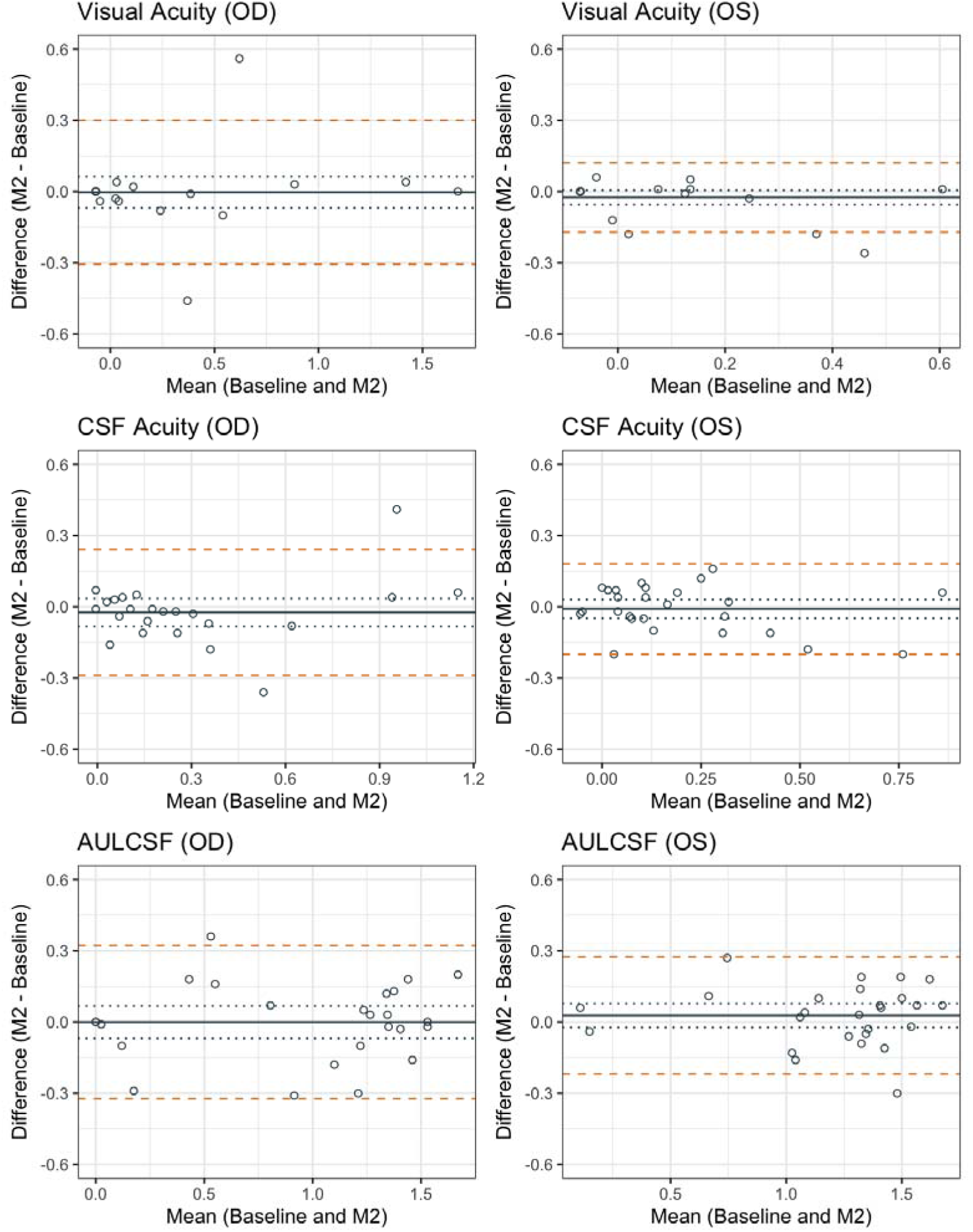
**Test-Retest Reliability of Chart-Based Vision Tests** Bland-Altman plots showing test–retest agreement for chart-based visual function parameters in the right eye (OD) and left eye (OS). Differences between baseline and second visit are plotted against the mean of the two measurements. The solid grey line represents the mean difference (bias), with dotted lines indicating the 95% confidence interval. The dashed orange lines show the 95% limits of agreement (LoA).

**Table 3.** Test-Retest Reliability of Chart-Based Vision Tests

|  | Mean difference<br>(95% CI) | SD of<br>differences | CoR<br>(95% CI) | Upper LoA<br>(95% CI) | Lower LoA<br>(95% CI) |
| --- | --- | --- | --- | --- | --- |
| **Visual Acuity**<br>**(RE, logMAR)** | -0.003 (-0.068, 0.062) | 0.155 | 0.303 (0.236; 0.425) | 0.300 (0.187; 0.413) | -0.306 (-0.419; -0.193) |
| **Visual Acuity**<br>**(LE, logMAR)** | -0.025 (-0.055, 0.005) | 0.074 | 0.146 (0.114; 0.201) | 0.121 (0.069; 0.173) | -0.170 (-0.223; -0.118) |
| **AULCSF (RE,**<br>**logCS\*logCPD)** | -0.0 (-0.068, 0.068) | 0.165 | 0.323 (0.252; 0.449) | 0.322 (0.205; 0.440) | -0.323 (-0.441; -0.205) |
| **AULCSF (LE,**<br>**logCS\*logCPD)** | 0.027 (-0.024, 0.078) | 0.126 | 0.247 (0.194; 0.341) | 0.274 (0.186; 0.362) | -0.219 (-0.308; -0.131) |
| **CSF Acuity**<br>**(RE, logMAR)** | -0.024 (-0.082, 0.035) | 0.135 | 0.265 (0.205; 0.374) | 0.241 (0.139; 0.342) | -0.288 (-0.390; -0.187) |
| **CSF Acuity**<br>**(LE, logMAR)** | -0.009 (-0.045, 0.030) | 0.098 | 0.191 (0.150; 0.264) | 0.182 (0.114; 0.250) | -0.200 (-0.269; -0.132) |

SD: standard deviation; CoR: coefficient of repeatability; LoA: limit of agreement; CI: confidence interval; RE: right eye; LE: left eye; logMAR: logarithm of the minimum angle of resolution.

### Retest-Reliability of Steady-State Microperimetry

Figure 3 shows the test-retest reliability of mesopic and scotopic microperimetry for mean sensitivity and pointwise sensitivity. Overall, the bias between test and retest was lowest in scotopic cyan microperimetry with a mean bias [95% CI] of 0.06 dB [-0.37, -0.49], followed by scotopic red (0.20 dB [-0.19, 0.60]) and mesopic microperimetry (0.36 dB [-0.06, 0.78]). Table 4 summarizes the results.

**Figure 3.**
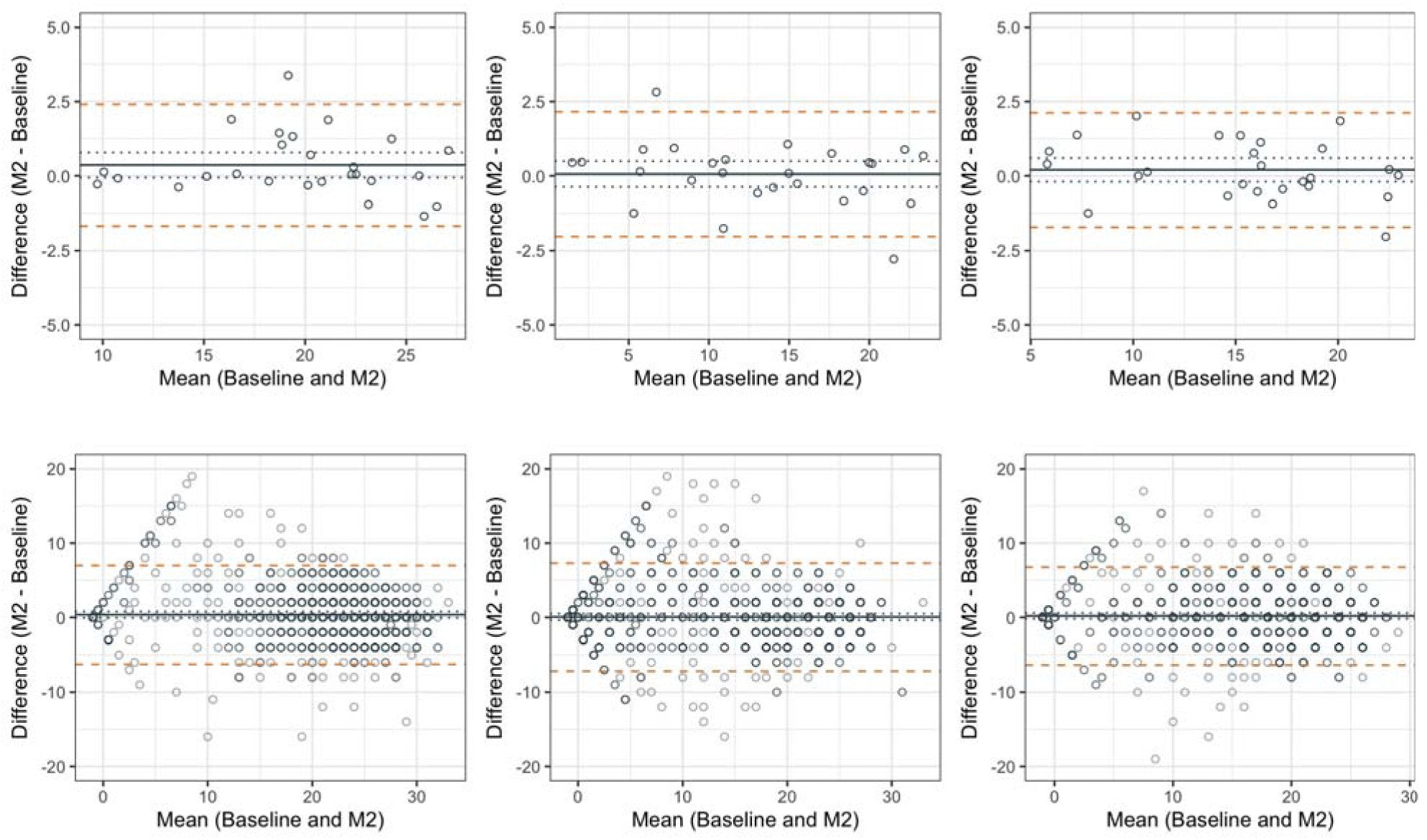
Test-Retest Reliability of Steady-State Microperimetry. Bland-Altman plots of the test-retest reliability of mesopic (left), scotopic cyan (center) and scotopic red (right) microperimetry. Mean sensitivity was reported in the first row, pointwise sensitivity in the second row. The solid grey indicates the average bias with a 95% CI (dotted lines). The dashed orange lines indicate the upper- and lower LoA. Data points in the second row are plotted in grey with transparency (alpha = 0.5); darker regions represent areas with higher data density due to overplotting. All values are shown in decibel (dB).

**Table 4.** Test-Retest Reliability in Steady-State Microperimetry

|  | Mean difference<br>(95% CI) | SD of<br>differences | CoR<br>(95% CI) | Upper LoA<br>(95% CI) | Lower LoA<br>(95% CI) |
| --- | --- | --- | --- | --- | --- |
| **Mean Sensitivity** |  |  |  |  |  |
| **Mesopic** | 0.36 (-0.06, 0.78) | 1.042 | 2.04 (1.44, 2.64) | 2.41 (1.80, 3.01) | -1.68 (-2.28, -1.07) |
| **Scotopic cyan** | 0.06 (-0.37, 0.49) | 1.068 | 2.09 (1.48, 2.71) | 2.16 (1.54, 2.77) | -2.03 (-2.65, -1.41) |
| **Scotopic red** | 0.20 (-0.19, 0.60) | 0.979 | 1.92 (1.35, 2.48) | 2.12 (1.55, 2.69) | -1.72 (-2.28, -1.15) |
| **Pointwise analysis** |  |  |  |  |  |
| **Mesopic** | 0.36 (-0.06, 0.78) | 3.377 | 6.62 (6.24, 7.00) | 6.98 (6.61, 7.36) | -6.26 (-6.63, -5.88) |
| **Scotopic cyan** | 0.06 (-0.37, 0.49) | 3.698 | 7.24 (6.86, 7.64) | 7.31 (6.92, 7.70) | -7.18 (-7.57, -6.80) |
| **Scotopic red** | 0.20 (-0.19, 0.60) | 3.349 | 6.56 (6.21, 6.92) | 6.77 (6.41, 7.12) | -6.36 (-6.71, -6.01) |

All values are shown in decibel (dB); |SD: standard deviation; CoR: coefficient of repeatability; LoA: limit of agreement; CI: confidence interval; RE: right eye; LE: left eye; logMAR: logarithm of the minimum angle of resolution.

For mean sensitivity, mesopic LoA ranged from -1.68 dB [-2.28, -1.07] to 2.41 dB [1.80, 3.01]. Scotopic cyan LoA ranged from a lower LoA of -2.03 dB [-2.65, -1.41] to an upper LoA of 2.16 dB [1.54, 2.77] while scotopic red mean sensitivity LoA ranged from a lower LoA of -1.72 dB [-2.28, - 1.15] to an upper LoA of 2.12 dB [1.55, 2.69] dB).

At a pointwise level, mesopic lower LoA was -6.26 dB [-6.63, -5.88] and upper LoA was 6.98 dB [6.61, 7.36]. Scotopic cyan LoA ranged from a lower LoA of -7.18 dB [-7.57, -6.80] to an upper LoA of 7.31 dB [6.92, 7.70]. The pointwise LoA of the scotopic red exams ranged from -6.36 dB [-6.71, - 6.01] to 6.77 [6.41, 7.12].

### Retest-Reliability of Dark-Adaptation Curve Parameters

Between the baseline exam and the retest-exam, no significant change in median cone rod break time (CRB), rod intercept time (RIT), S2 slope, cone threshold, final rod threshold, initial threshold, and exponential cone recovery time constant was observed between the baseline and second visits for the study eye (all p-values > 0.05; Table 5). These findings indicate that dark-adaptation curve parameters remained stable between the two measurements.

**Table 5.**
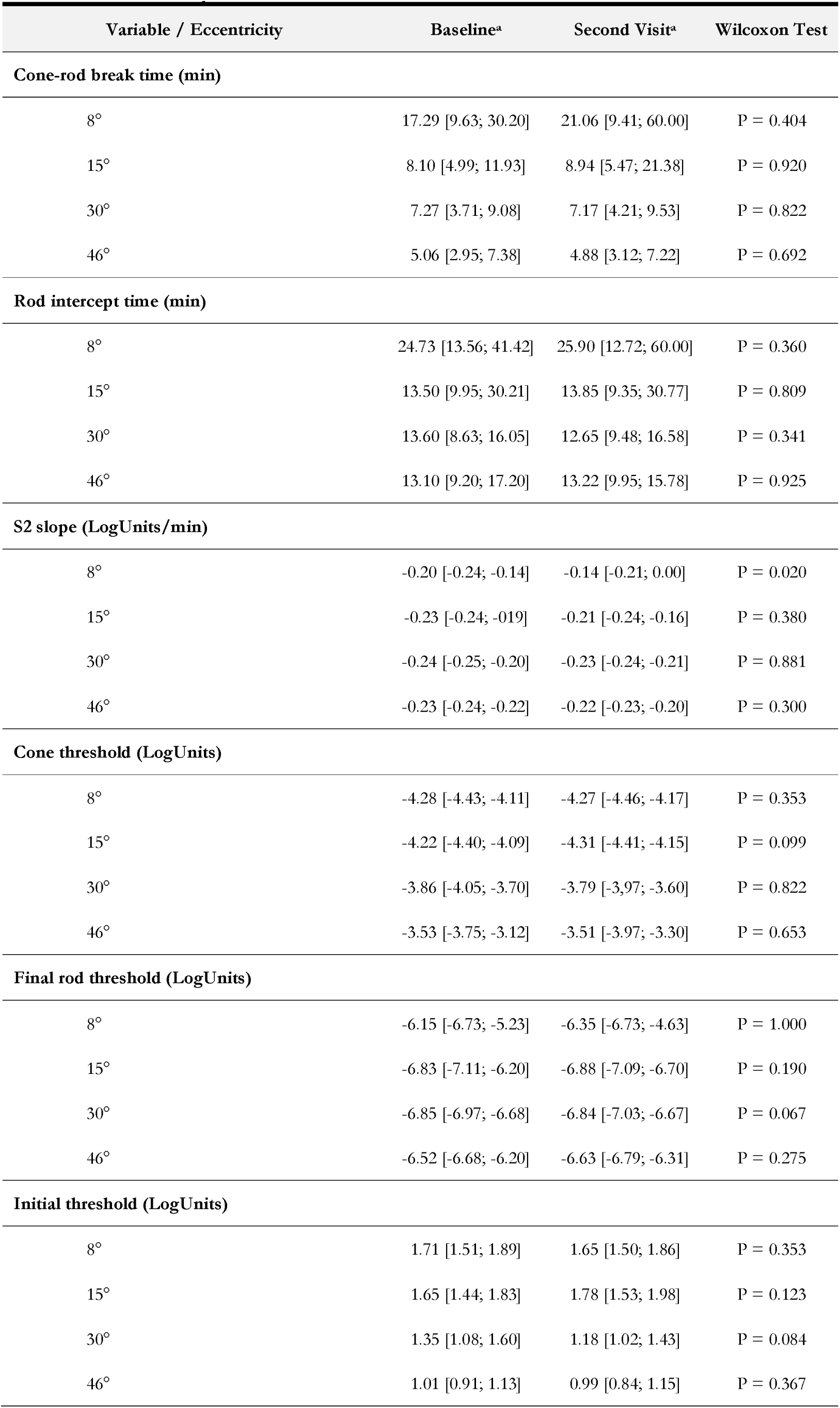
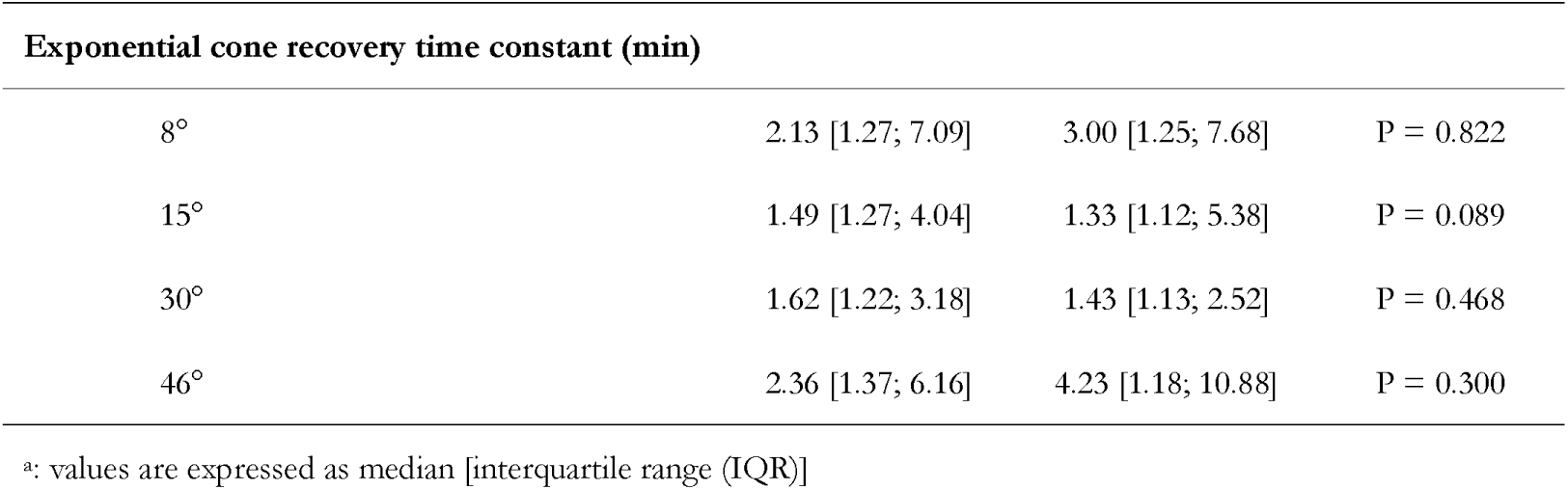
Dark-Adaptation Curve Parameters

| Variable / Eccentricity | Baseline<sup>a</sup> | Second Visit<sup>a</sup> | Wilcoxon Test |
| --- | --- | --- | --- |
| **Cone-rod break time (min)** |  |  |  |
| 8° | 17.29 [9.63; 30.20] | 21.06 [9.41; 60.00] | P = 0.404 |
| 15° | 8.10 [4.99; 11.93] | 8.94 [5.47; 21.38] | P = 0.920 |
| 30° | 7.27 [3.71; 9.08] | 7.17 [4.21; 9.53] | P = 0.822 |
| 46° | 5.06 [2.95; 7.38] | 4.88 [3.12; 7.22] | P = 0.692 |
| **Rod intercept time (min)** |  |  |  |
| 8° | 24.73 [13.56; 41.42] | 25.90 [12.72; 60.00] | P = 0.360 |
| 15° | 13.50 [9.95; 30.21] | 13.85 [9.35; 30.77] | P = 0.809 |
| 30° | 13.60 [8.63; 16.05] | 12.65 [9.48; 16.58] | P = 0.341 |
| 46° | 13.10 [9.20; 17.20] | 13.22 [9.95; 15.78] | P = 0.925 |
| **S2 slope (LogUnits/min)** |  |  |  |
| 8° | -0.20 [-0.24; -0.14] | -0.14 [-0.21; 0.00] | P = 0.020 |
| 15° | -0.23 [-0.24; -0.19] | -0.21 [-0.24; -0.16] | P = 0.380 |
| 30° | -0.24 [-0.25; -0.20] | -0.23 [-0.24; -0.21] | P = 0.881 |
| 46° | -0.23 [-0.24; -0.22] | -0.22 [-0.23; -0.20] | P = 0.300 |
| **Cone threshold (LogUnits)** |  |  |  |
| 8° | -4.28 [-4.43; -4.11] | -4.27 [-4.46; -4.17] | P = 0.353 |
| 15° | -4.22 [-4.40; -4.09] | -4.31 [-4.41; -4.15] | P = 0.099 |
| 30° | -3.86 [-4.05; -3.70] | -3.79 [-3.97; -3.60] | P = 0.822 |
| 46° | -3.53 [-3.75; -3.12] | -3.51 [-3.97; -3.30] | P = 0.653 |
| **Final rod threshold (LogUnits)** |  |  |  |
| 8° | -6.15 [-6.73; -5.23] | -6.35 [-6.73; -4.63] | P = 1.000 |
| 15° | -6.83 [-7.11; -6.20] | -6.88 [-7.09; -6.70] | P = 0.190 |
| 30° | -6.85 [-6.97; -6.68] | -6.84 [-7.03; -6.67] | P = 0.067 |
| 46° | -6.52 [-6.68; -6.20] | -6.63 [-6.79; -6.31] | P = 0.275 |
| **Initial threshold (LogUnits)** |  |  |  |
| 8° | 1.71 [1.51; 1.89] | 1.65 [1.50; 1.86] | P = 0.353 |
| 15° | 1.65 [1.44; 1.83] | 1.78 [1.53; 1.98] | P = 0.123 |
| 30° | 1.35 [1.08; 1.60] | 1.18 [1.02; 1.43] | P = 0.084 |
| 46° | 1.01 [0.91; 1.13] | 0.99 [0.84; 1.15] | P = 0.367 |

| **Exponential cone recovery time constant (min)** |  |  |  |
| --- | --- | --- | --- |
| 8° | 2.13 [1.27; 7.09] | 3.00 [1.25; 7.68] | P = 0.822 |
| 15° | 1.49 [1.27; 4.04] | 1.33 [1.12; 5.38] | P = 0.089 |
| 30° | 1.62 [1.22; 3.18] | 1.43 [1.13; 2.52] | P = 0.468 |
| 46° | 2.36 [1.37; 6.16] | 4.23 [1.18; 10.88] | P = 0.300 |

<sup>a</sup>: values are expressed as median [interquartile range (IQR)]

Test–retest reliability was assessed for each dark adaptation parameter by calculating the mean difference, standard deviation of differences, coefficient of repeatability (CoR), and 95% limits of agreement (LoA), including corresponding confidence intervals. The results are summarized in Table 6. The Bland-Altman plots for RIT and final rod threshold are shown in Figure 4. Bland-Altman plots for all dark adaptation curve parameters are provided in Supplementary Figure S2.

**Figure 4.**
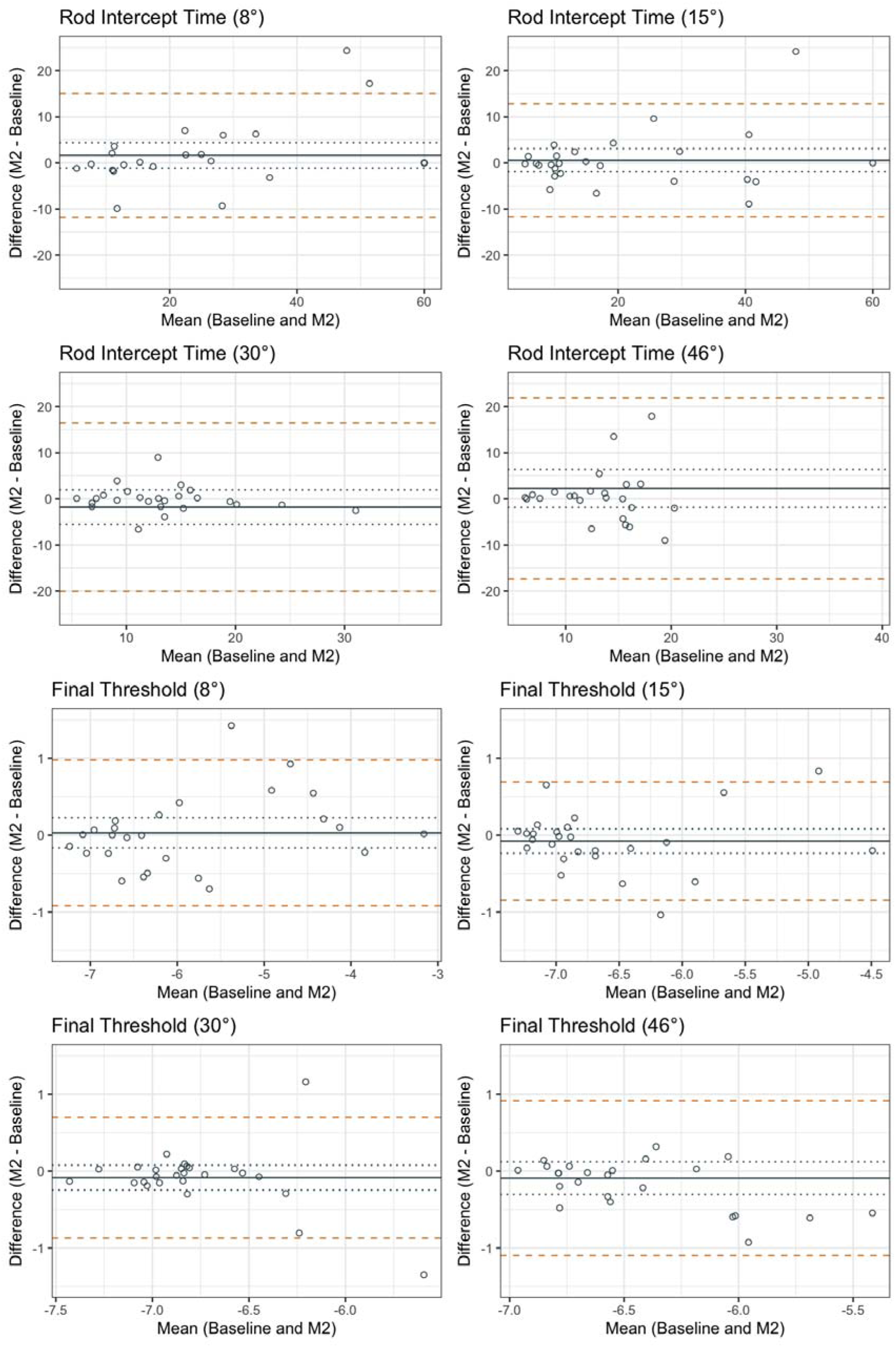
Test-Retest Reliability of Dark-Adaptation Curve Parameters Bland-Altman plots for rod intercept time (top panels) and final rod threshold (bottom panels) at four retinal eccentricities (8°, 15°, 30°, and 46°). Each plot displays the difference between test and retest measurements (M2 – Baseline) against their mean. The solid grey line represents the mean difference (bias), with dotted lines indicating the 95% confidence interval of the bias. The orange dashed lines show the 95% limits of agreement (LoA).

**Table 6.** Test-Retest Reliability of Dark-Adaptation Curve Parametersw

| Parameter/ Eccentricity | Mean difference | SD of differences | CoR (CI 95%) | Upper LoA (CI 95%) | Lower LoA (CI 95%) |
| --- | --- | --- | --- | --- | --- |
| **Cone rod break time (min)** |  |  |  |  |  |
| 8° | 3.782 | 13.058 | 25.593 (20.071; 35.329) | 29.375 (20.243; 38.506) | -21.811 (-30.943; -12.680) |
| 15° | 2.134 | 12.62 | 24.735 (19.398; 34.144) | 26.869 (18.044; 35.694) | -22.600 (-31.426; -13.775) |
| 30° | 0.313 | 2.623 | 5.141 (4.032; 7.097) | 5.454 (3.620; 7.289) | -4.828 (-6.662; -2.994) |
| 46° | 2.419 | 10.942 | 21.447 (16.747; 29.836) | 23.866 (16.041; 31.690) | -19.028 (-26.853; -11.204) |
| **Rod intercept time (min)** |  |  |  |  |  |
| 8° | 1.625 | 6.844 | 13.414 (10.520; 18.516) | 15.039 (10.253; 19.824) | -11.789 (-16.574; -7.003) |
| 15° | 0.569 | 6.236 | 12.222 (9.585; 16.871) | 12.791 (8.430; 17.152) | -11.653 (-16.013; -7.292) |
| 30° | -1.829 | 9.31 | 18.247 (14.311; 25.189) | 16.419 (9.908; 22.929) | -20.076 (-26.587; -13.566) |
| 46° | 2.254 | 10.0 | 19.599 (15.304; 27.265) | 21.853 (14.703; 29.003) | -17.345 (-24.495; -10.195) |
| **S2 slope (LogUnits/min)** |  |  |  |  |  |
| 8° | 0.041 | 0.083 | 0.162 (0.127; 0.224) | 0.203 (0.146; 0.261) | -0.121 (-0.179; -0.063) |
| 15° | 0.016 | 0.063 | 0.123 (0.097; 0.170) | 0.139 (0.095; 0.183) | -0.107 (-0.151; -0.063) |
| 30° | 0.001 | 0.054 | 0.105 (0.083; 0.145) | 0.107 (0.069; 0.144) | -0.104 (-0.141; -0.066) |
| 46° | 0.017 | 0.05 | 0.099 (0.077; 0.137) | 0.116 (0.080; 0.152) | -0.081 (-0.117; -0.045) |
| **Cone threshold (LogUnits)** |  |  |  |  |  |
| 8° | -0.055 | 0.263 | 0.515 (0.404; 0.710) | 0.459 (0.276; 0.643) | -0.570 (-0.753; -0.386) |
| 15° | -0.102 | 0.288 | 0.565 (0.443; 0.780) | 0.462 (0.261; 0.664) | -0.667 (-0.869; -0.466) |
| 30° | 0.062 | 0.359 | 0.705 (0.553; 0.973) | 0.766 (0.515; 1.018) | -0.643 (-0.894; -0.391) |
| 46° | -0.095 | 0.329 | 0.645 (0.504; 0.898) | 0.551 (0.315; 0.786) | -0.740 (-0.975; -0.504) |
| **Final rod threshold (LogUnits)** |  |  |  |  |  |
| 8° | 0.03 | 0.483 | 0.946 (0.742; 1.306) | 0.977 (0.639; 1.315) | -0.916 (-1.254; -0.578) |
| 15° | -0.077 | 0.393 | 0.770 (0.604; 1.063) | 0.693 (0.418; 0.968) | -0.847 (-1.122; -0.572) |
| 30° | -0.085 | 0.402 | 0.787 (0.617; 1.086) | 0.702 (0.422; 0.983) | -0.872 (-1.153; -0.591) |
| 46° | -0.091 | 0.514 | 1.008 (0.787; 1.402) | 0.917 (0.549; 1.285) | -1.099 (-1.467; -0.731) |
| **Initial threshold (LogUnits)** |  |  |  |  |  |
| 8° | -0.213 | 0.764 | 1.497 (1.174; 2.067) | 1.284 (0.750; 1.818) | -1.711 (-2.245; -1.176) |
| 15° | 0.118 | 0.442 | 0.867 (0.680; 1.196) | 0.985 (0.676; 1.294) | -0.748 (-1.058; -0.439) |
| 30° | -0.165 | 0.392 | 0.769 (0.603; 1.061) | 0.603 (0.329; 0.877) | -0.934 (-1.208; -0.660) |
| 46° | -0.008 | 0.192 | 0.375 (0.293; 0.522) | 0.368 (0.231; 0.505) | -0.383 (-0.520; -0.246) |
| **Exponential cone recovery time constant (min)** |  |  |  |  |  |
| 8° | 0.338 | 3.891 | 7.627 (5.982; 10.529) | 7.966 (5.244; 10.687) | -7.289 (-10.010; -4.567) |
| 15° | -0.294 | 4.484 | 8.789 (6.893; 12.132) | 8.495 (5.359; 11.630) | -9.083 (-12.219; -5.948) |
| 30° | -1.357 | 6.236 | 12.223 (9.586; 16.873) | 10.866 (6.504; 15.227) | -13.581 (-17.942; -9.219) |
| 46° | 1.396 | 5.266 | 10.321 (8.059; 14.357) | 11.717 (7.952; 15.482) | -8.924 (-12.689; -5.159) |

SD: standard deviation; CoR: coefficient of repeatability; LoA: limit of agreement; CI: confidence interval; logMAR: logarithm of the minimum angle of resolution

## DISCUSSION

Reliable functional endpoints are essential for evaluating treatment efficacy in rare diseases such as PXE, particularly in the context of emerging interventional trials. If validated, functional ophthalmologic measures in PXE may offer clear advantages over currently employed primary endpoints—such as low-dose computed tomography of the legs and carotid arteries (e.g., ClinicalTrials.gov ID NCT05832580)—in terms of both actual patient relevance and feasibility, as well as image quality, cost, and patient safety. Despite their potential, to our knowledge, no prior study has systematically assessed the test-retest reliability of functional outcome measures in PXE.

In this study, we evaluated a broad array of functional assessments. BCVA exhibited high test-retest variability, with limits of agreement ranging from -0.31 to 0.30 logMAR. However, BCVA often fails to capture the full extent of visual dysfunction, particularly in the presence of secondary complications such as MNVs or atrophic changes outside of the fovea—features commonly observed in late stages of PXE. Similarly, other chart-based assessments, such as contrast sensitivity, are likely driven mostly by foveal and parafoveal changes. Thus, like AMD, other tests to determine visual functions are explored.^36^

Consistent with previously published test-retest studies—particularly those employing the same MAIA microperimetry device—our microperimetry results demonstrated reliability metrics comparable to those reported in patients with AMD and other macular diseases.^25–30,35^ Our pointwise limits of agreements of ±6.8 dB fall well between the published data of healthy volunteers and patients with retinal diseases, with LoAs typically ranging from ± 3.4 dB to 9.5 dB (Supplementary Table S1). Notably, we observed comparable test-retest reliability for dark-adapted scotopic red and cyan stimuli, test settings for which systematic reliability data are currently sparse. These findings suggest that scotopic microperimetry may offer a reliable means of detecting early retinal dysfunction, potentially complementing conventional mesopic assessments.

In contrast, our dark adaptation results—measured by rod intercept time —differed from previously published data.^15,21,29^ Of note, our cohort had more advanced disease than in prior studies. Earlier test–retest data are mostly from healthy subjects or patients with early or intermediate AMD.^15,21–35^ In contrast, several of our patients reached the ceiling of the test (RIT >60 minutes), underscoring severity. Markedly longer test durations, fatigue, and fixation instability likely contributed to increased variability. Currently, fundus-controlled dark adaptometry methods are in development aiming to improve repeatability.^37^

In the main analysis, non-measurable RITs were imputed as 60 minutes. This yielded conservative limits of agreement (LOAs), which were heavily influenced by a few patients with markedly delayed adaptation. In clinical trials, such patients (those near or beyond the maximum test window) would be excluded or studied with extended protocols. Accordingly, we also evaluated how test–retest reliability improves with exclusion of these patients (Supplementary Figure S3 and Supplementary Table S2). Following outlier removal (eight patients at 8° eccentricity, two at 15°, one at 30°, and one at 46°), reproducibility improved, and results aligned with values reported in other retinal diseases (Supplementary Table S1).

Beyond their utility in regulatory and research frameworks, functional measures such as mesopic and scotopic microperimetry also offer advantages in capturing aspects of visual performance that are directly relevant to patients’ daily experiences. Conventional imaging endpoints may overlook subtle but meaningful impairments—such as difficulties with night vision or visual contrast—that significantly affect quality of life and occur early in the patient’s journey^12^.

In summary, our findings demonstrate that mesopic and scotopic microperimetry have a robust test-retest reliability, aligning with prior data from AMD and other macular diseases, and showing potential as reliable functional endpoints in PXE. Although dark adaptometry showed higher variability—likely due to fixation instability and advanced disease features—its refinement through fundus-controlled approaches may enhance its utility in future studies.

## Data Availability

All data produced in the present study are available upon reasonable request to the authors

