## Supplementary Material for "Retest-Reliability of Cone and Rod Function Assessments in Pseudoxanthoma elasticum: PROPXE Study Report 3"

### **Supplementary Figure S1. Perimetry Grid**

The dot plot shows the perimetry grid included in this study. The color indicates the median mesopic sensitivity. Right eyes and left eyes were harmonized, with negative values (-15°) corresponding to temporal and positive values (+15°) to nasal locations. The '0 dB' area on the nasal side overlaps with the optic nerve head.

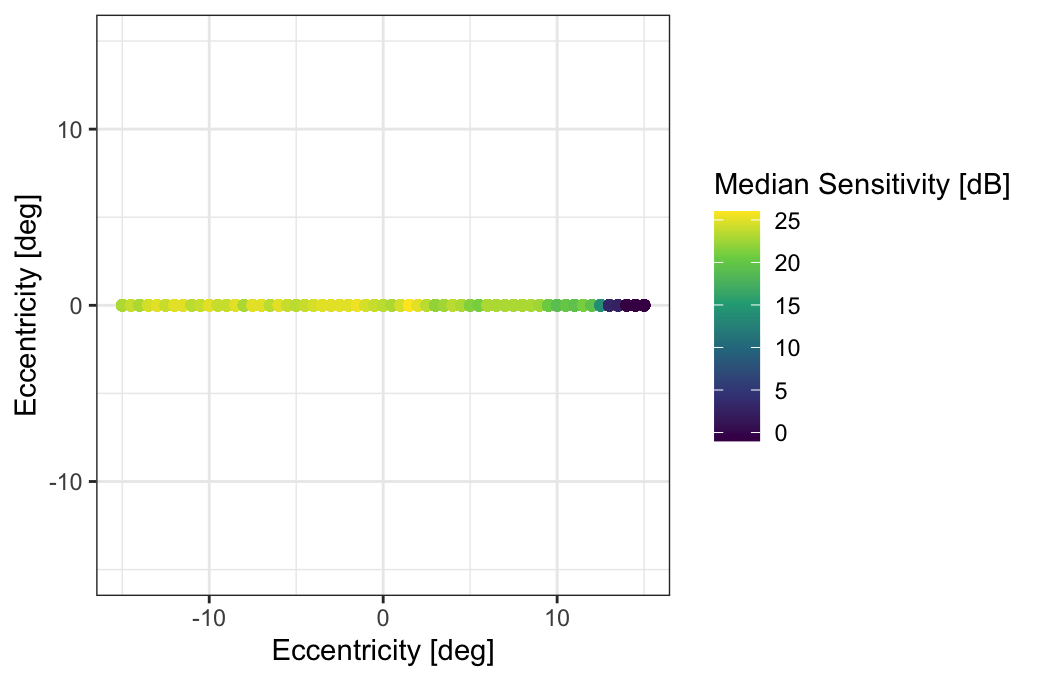

### **Supplementary Figure S2. Test-Retest Reliability of Dark-Adaptation Curve Parameters**

Bland-Altman plots for all dark-adaptation curve parameters (cone-rod break time, cone threshold, rod intercept time, S2 slope, initial threshold, exponential cone recovery time constant, final rod threshold) at four retinal eccentricities (8°, 15°, 30°, and 46°). Each plot displays the difference between test and retest measurements (M2 – Baseline) against their mean. The solid grey line represents the mean difference (bias), with dotted lines indicating the 95% confidence interval of the bias. The orange dashed lines show the 95% limits of agreement (LoA).

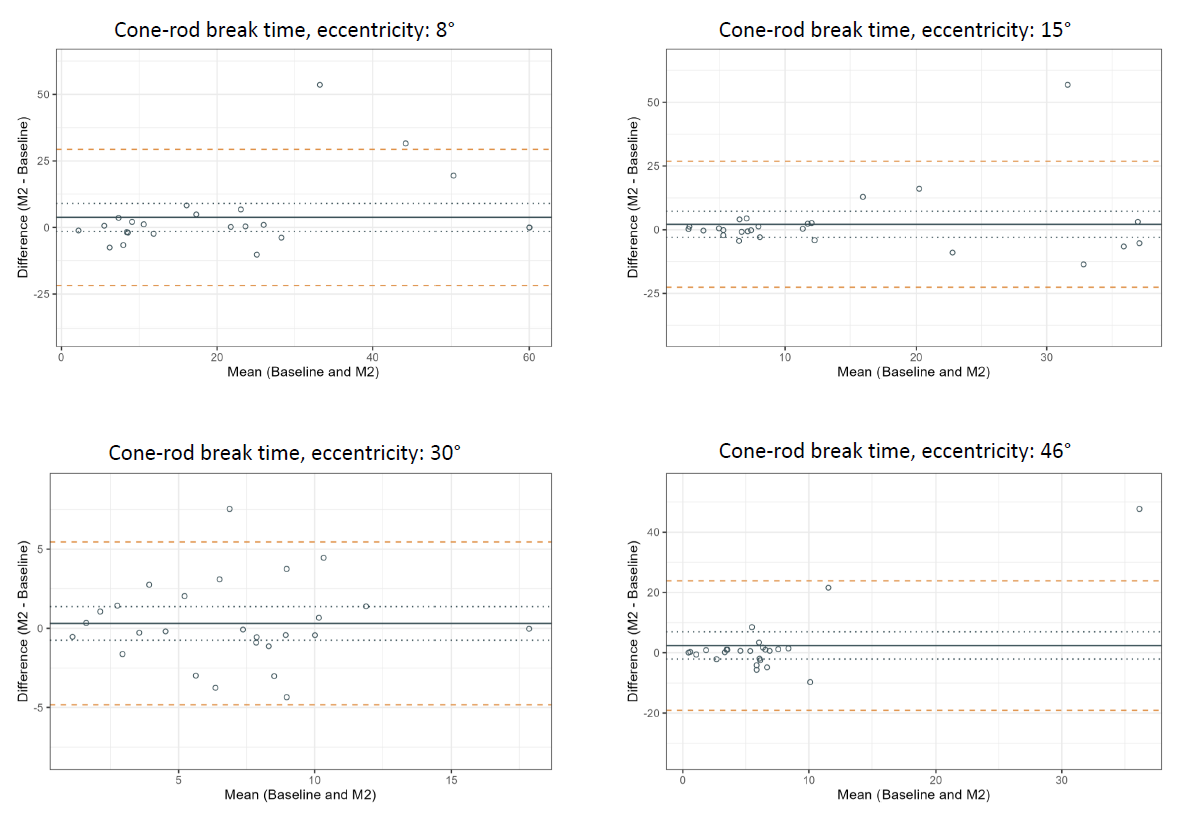

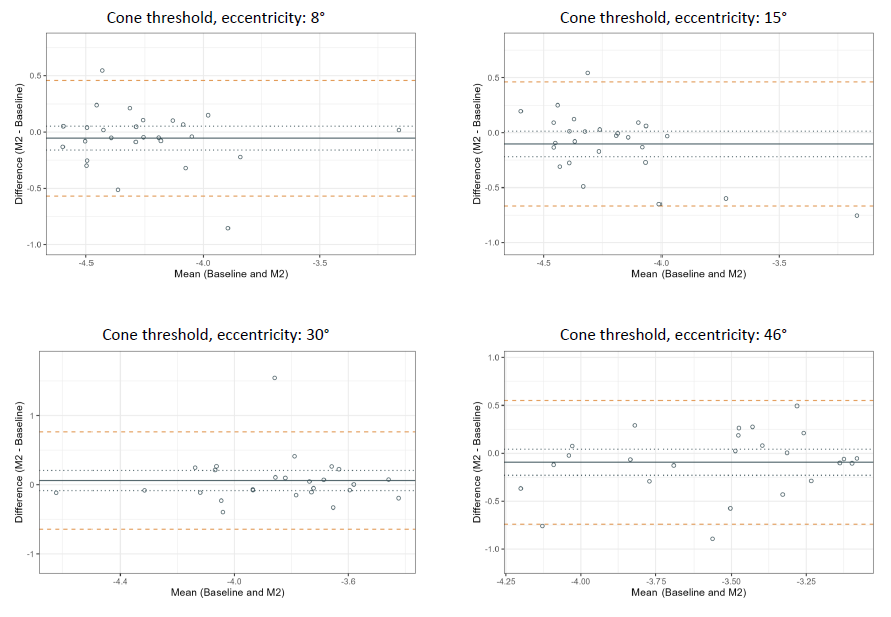

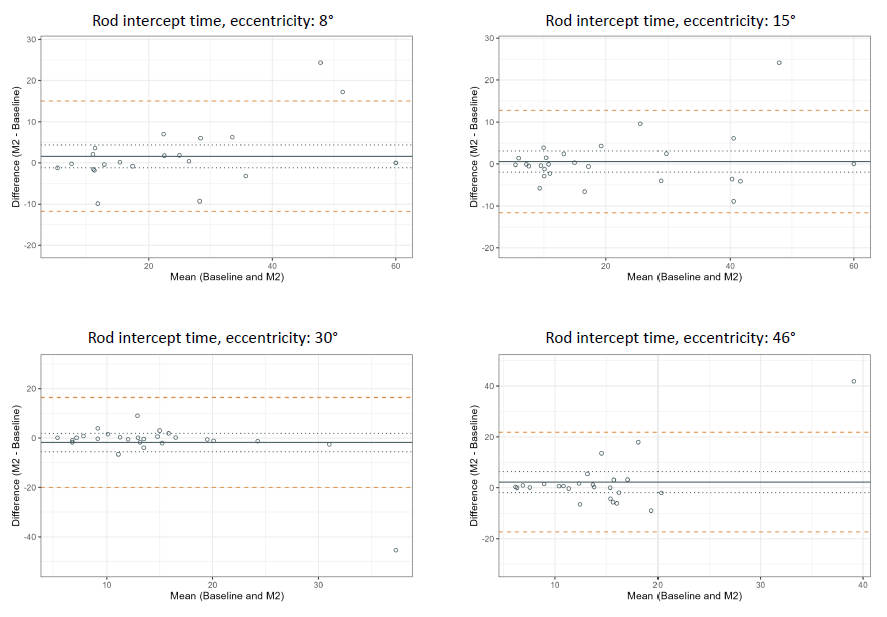

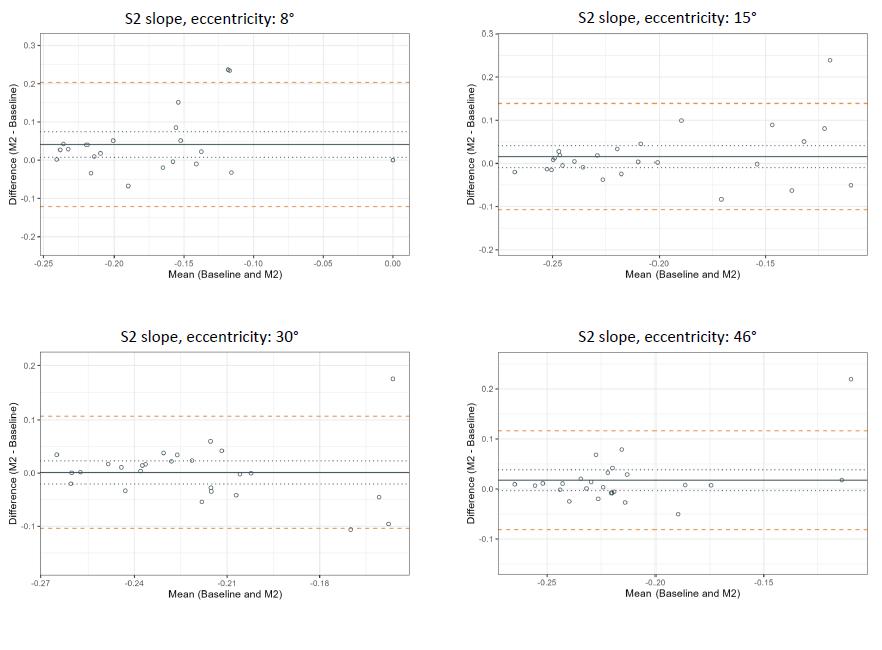

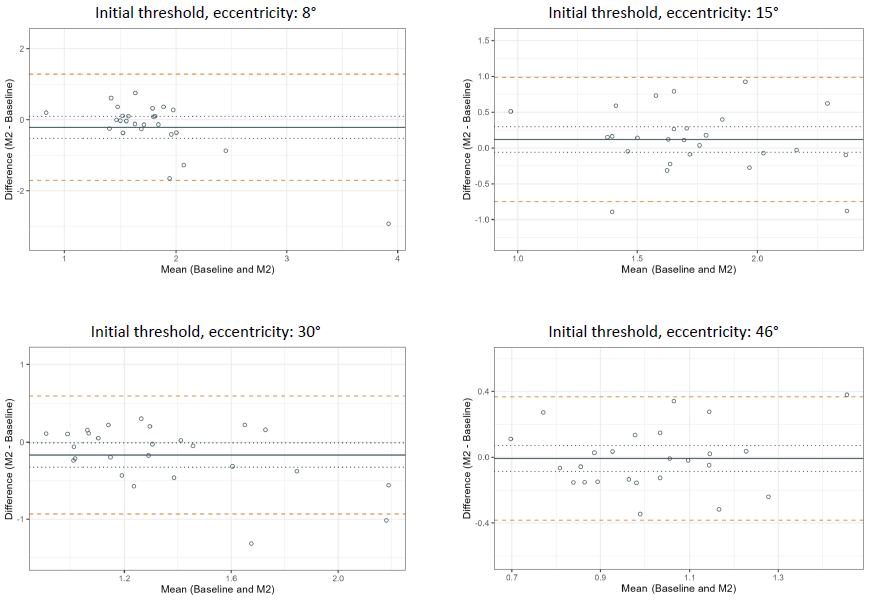

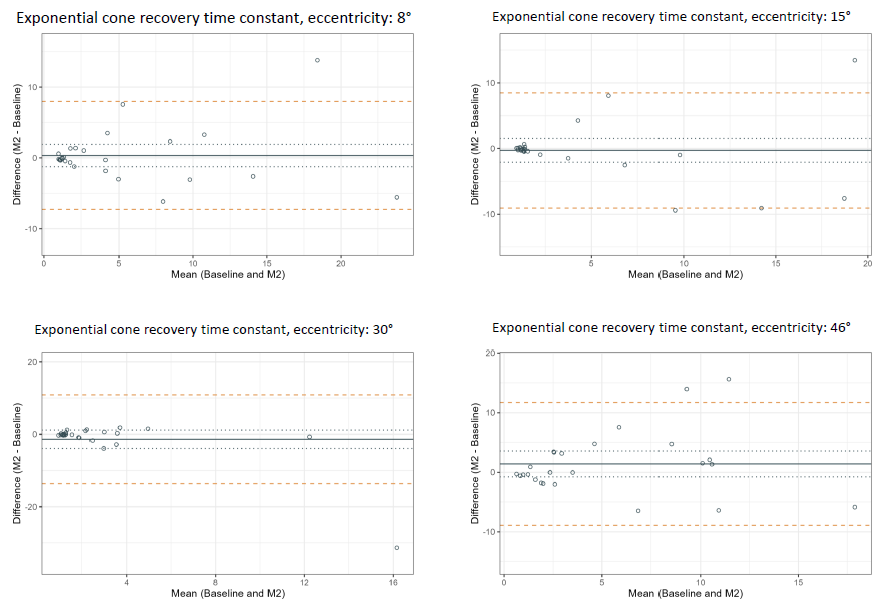

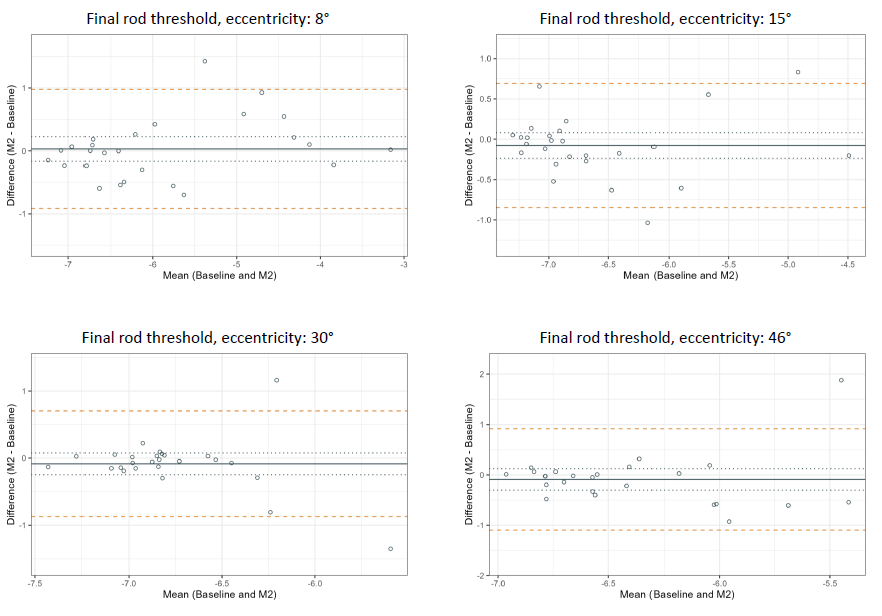

### **Supplementary Figure S3. Bland Altman Plots of Rod Intercept Time (RIT), after Exclusion of Eyes with a RIT >60 min**

Each plot shows the agreement between baseline and follow-up measurements for individual eccentricities (8°, 15°, 30°, and 46°). Outliers corresponding to non-measurable or markedly delayed dark adaptation (>60 min) were excluded (eight eyes at 8°, two at 15°, one at 30°, and one at 46°). The solid grey line represents the mean difference (bias), with dotted lines indicating the 95% confidence interval of the bias. The orange dashed lines show the 95% limits of agreement (LoA).

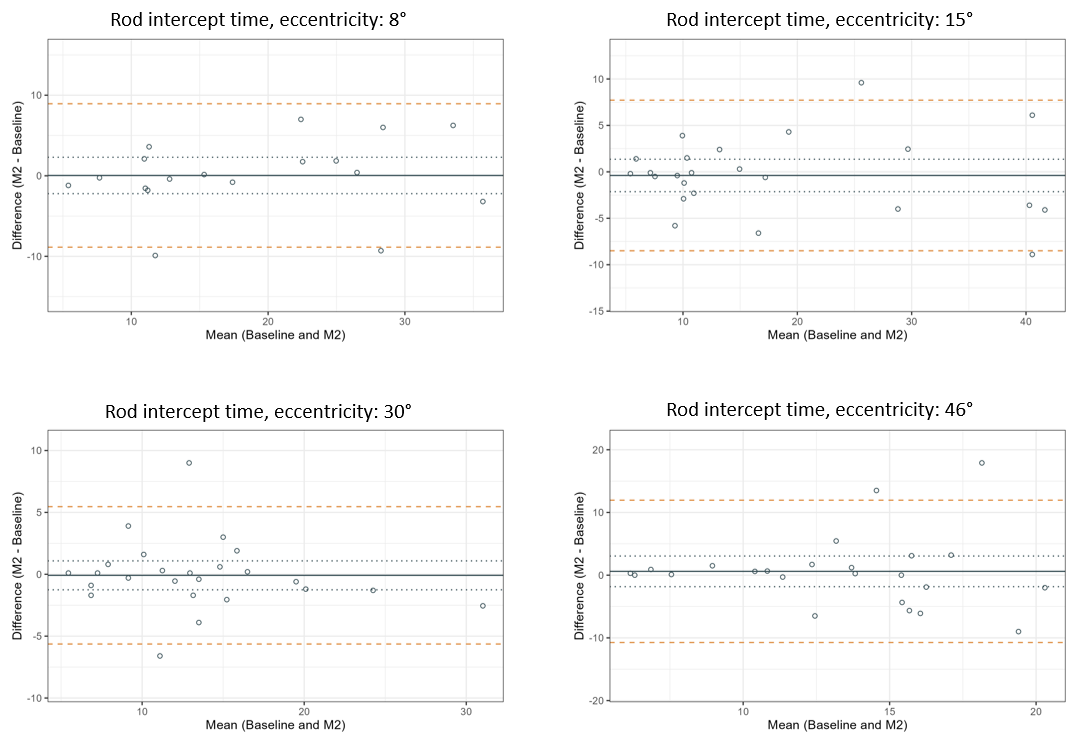

### **Supplementary Figure S4. Image Analysis of Pseudoxanthoma Elasticum (PXE) Characteristics**

Panel A shows a nine-gaze 55° infrared (IR) image collage of an exemplary eye with PXE. Panel B shows the Peau d’orange (PdO, upper half) and subretinal drusenoid deposits (SDD, bottom half). The dashed white lines delineate the area of continuously calcified Bruch’s membrane ('Coquille d'œuf'). Panel C shows an angioid streak highlighted in red, not exceeding the Coquille d'œuf.

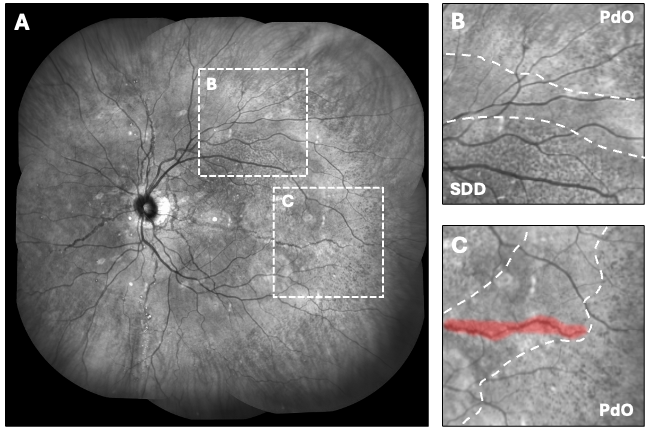

### **Supplementary Figure S5. Comparison of Temporal Peau d’Orange Extend to Previously Published Data**

The two dot plots show the temporal inner (left plot) and temporal outer (right plot) Peau d’orange boundary extents measured in our study (yellow/gold dots) and in a Dutch cohort.^3^ The latter data were derived from Figure 3 in Risseeuw et al. 2021.^3^

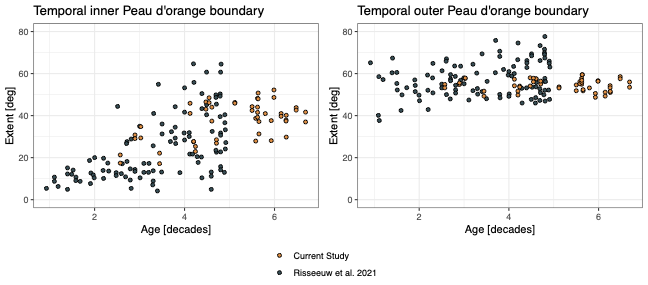

### **Supplementary Figure S6. Association Between the Temporal Inner Peau d’Orange Boundary and Delayed Dark Adaptation**

The panels show the relationship between the temporal inner Peau d’orange boundary and the rod-intercept time (RIT) as a measure of dark adaptation at 8°, 15°, and 30° eccentricity. The regression lines for the associations between the temporal inner boundary extent and RIT at 8° and 15°, respectively, were derived using a non-linear mixed model with a step function.

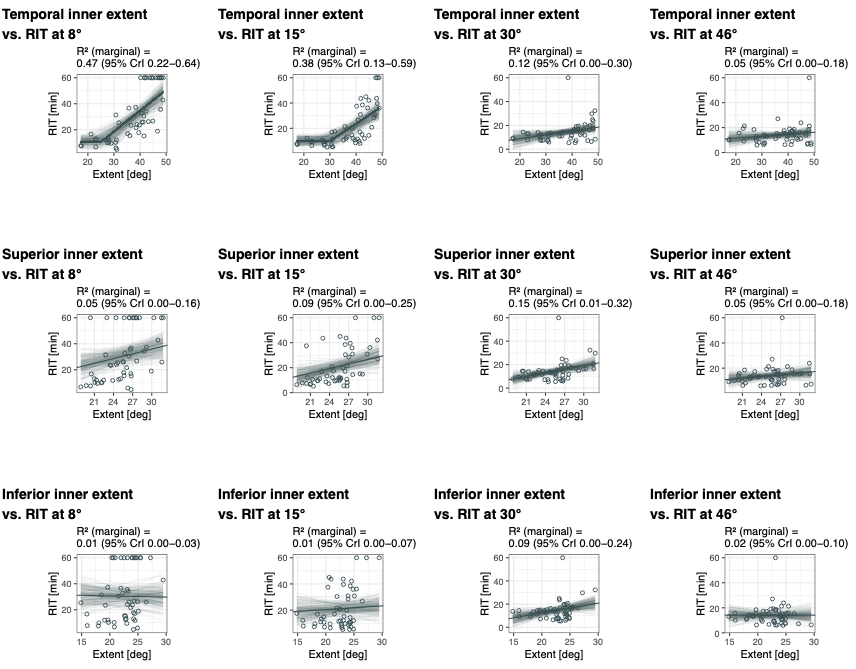

### **Supplementary Table S1. Literature Review of Retest-Reliability of Dark Adaptometry and Dark-Adapted Perimetry Testing**

The following literature search was performed with the search queries: 'test-retest variability AND microperimetry', 'test-retest repeatability AND microperimetry', 'test-retest repeatability AND dark-adapted perimetry', 'test-retest variability AND dark-adapted perimetry'.

**Supplementary Table S1A. Light- and Dark-Adapted (Free-Viewing) Perimetry**

| **First Author** | **Year** | **PMID** | **Perimeter** | **Study population** | **N (eyes; patients)** | **Age (mean ± SD [range])** | **BCVA** | **LA Orange-on-White [dB]** | **DA Cyan [dB]** | **DA Red [dB]** |
| --- | --- | --- | --- | --- | --- | --- | --- | --- | --- | --- |
| Uddin | 2020 | 32832236 | Medmont DACP | HV, iAMD | 12; 12 | 80 [63–90] | ≥ 20/63 | / | ± 7.2 | ± 5.9 |
| Tan | 2018 | 29736324 | Medmont DACP | intermediate/late dAMD | 7; 7 | 71 ± 6 [62–81] | ≥ 20/60 | / | ± 11.7 | ± 8.4 |
| Bennett | 2019 | 30901388 | Medmont DACP | IRD | 14; 14 | 40 ± 15 | ≥ 20/500 | / | ± 8.5 (intra-sessions), ± 9.8 (inter-session) | / |
| Cideciyan | 2018 | 30208424 | Modified HFA | XLRP | 26; 13 | 29 ± 7 [18–42] | / | ± 5.1 | ± 9.6 | / |

**Supplementary Table S1B. Light- and Dark-Adapted Microperimetry**

| **First Author** | **Year** | **PMID** | **Perimeter** | **Study population** | **N(eyes; patients)** | **Age (mean ± SD [range])** | **BCVA** | **Mesopic W-on-W [dB]** | **DA Cyan [dB]** | **DA Red [dB]** |
| --- | --- | --- | --- | --- | --- | --- | --- | --- | --- | --- |
| Pfau | 2017 | 27997924 | S-MAIA | HV | 30; 30 | 34 [13–80] | ≥ 20/25 | [–5.08, 4.29] (PWS) | [–4.72, 5.65] | / |
| Pfau | 2017 | 28692722 | S-MAIA | Retinal diseases | 52; 52 | 62 ± 17 [19–90] | / | ± 5.79 (PWS) | ± 4.72 | / |
| Pfau | 2020 | 30300264 | S-MAIA | GA secondary to AMD | 25; 25 | 77 ± 7 [64–90] | 20/68 | ± 6.64 (PWS) | ± 5.78 | / |
| Welker | 2018 | 30372731 | S-MAIA | iAMD | 23; 23 | 67 ± 8 [50–85] | ≥ 20/50 | ± 4.4 (PWS) | / | ± 4.52 |

**Supplementary Table S1C. Dark-Adaptometry**

| **First Author** | **Year** | **PMID** | **Perimeter** | **Study population** | **N (eyes; patients)** | **Age (mean ± SD [range])** | **BCVA** | **Cone Tau** | **Cone Threshold [dB]** | **Cone-Rod Break [min]** |
| --- | --- | --- | --- | --- | --- | --- | --- | --- | --- | --- |
| Oertli, | 2023 | 38112496 | S-MAIA | HV | 10; 10 | 32 [IQR 27–58] | –0.07 logMAR | / | [–2.8, 3.3] | [–3.94, 2.78] |
| Uddin | 2020 | 32832236 | Medmont DACP | HV, iAMD | 12; 12 | 80 [63–90] | ≥ 20/63 | / | ± 3.9 | / |
| Higgins | 2023 | 37477933 | AdaptDx | early AMD / intermediate AMD / late AMD | variable | 72 ± 6 / 71 ± 8 / 75 ± 6 | 0.01 logMAR / 0.02 logMAR / 0.78 logMAR | / | / | / |

**Supplementary Table S1C Continued**

| **First Author** | **Year** | **PMID** | **Perimeter** | **Rod Intercept Time [min]** | **Rate of Rod Decay / S2 [decades/min]** | **Final Threshold [dB]** |
| --- | --- | --- | --- | --- | --- | --- |
| Oertli | 2023 | 38112496 | S-MAIA | [–4.55, 3.11] | [–0.03, 0.03] | [–3.4, 2.8] |
| Uddin | 2020 | 32832236 | Medmont DACP | ± 7.6 | ± 0.075 | ± 6.6 (superior retina); ± 3.5 (inferior retina) |
| Higgins. | 2023 | 37477933 | AdaptDx | early AMD [–2.24, 2.33], N = 26 | int. AMD [–3.03, 3.43], N = 103 | late AMD [–1.77, 1.56], N = 8 |

### **Supplementary Table S2. Test-Retest Reliability of Rod Intercept Time (RIT), after** **Exclusion of Eyes with a RIT >60 min**

This analysis assesses the reproducibility of RIT measurements after removing extreme outliers corresponding to eyes with non-measurable or markedly delayed dark adaptation (>60 min). Eight eyes at 8° eccentricity, two at 15°, one at 30°, and one at 46° were excluded. Mean differences, standard deviations of differences, coefficients of repeatability (CoR), and 95% confidence intervals (CIs) for Bland–Altman limits of agreement (LoA) are shown for each eccentricity.

| **Eccentricity** | **Mean difference** | **SD of differences** | **CoR (CI 95%)** | **Upper LoA (CI 95%)** | **Lower LoA (CI 95%)** |
| --- | --- | --- | --- | --- | --- |
| 8° | 0.039 | 4.542 | 8.903 (6.681; 13.347) | 8.942 (5.007; 12.876) | -8.864 (-12.799; -4.930) |
| 15° | -0.390 | 4.137 | 8.109 (6.302; 11.374) | 7.719 (4.691; 10.747) | -8.498 (-11.526; -5.470) |
| 30° | -0.086 | 2.832 | 5.550 (4.334; 7.721) | 5.464 (3.439; 7.489) | -5.636 (-7.661; -3.611) |
| 46° | 0.606 | 5.789 | 11.346 (8.818; 15.916) | 11.952 (7.715; 16.189) | -10.740 (-14.977; -6.503) |

### **Supplementary Table S3. Inter-Reader Reliability of Inner- and Outer Peau d’Orange Boundary**

| **Visit** | **Meridian** | **Boundary** | **Standard Error of Measurement** | **Coefficient of Variation** | **Intraclass Correlation Coefficient (two-way random, agreement)** |
| --- | --- | --- | --- | --- | --- |
| V1 | Temporal | Inner | 2.68 [2.11, 3.17] | 0.07 [0.06, 0.09] | 0.92 [0.89, 0.95] |
|  |  | Outer | 2.50 [1.36, 3.68] | 0.05 [0.02, 0.07] | 0.48 [0.30, 0.64] |
| V1 | Superior | Inner | 2.35 [1.86, 2.87] | 0.09 [0.07, 0.11] | 0.66 [0.54, 0.76] |
|  |  | Outer | 2.22 [1.88, 2.49] | 0.06 [0.05, 0.07] | 0.51 [0.32, 0.66] |
| V1 | Inferior | Inner | 2.93 [2.32, 3.47] | 0.13 [0.11, 0.16] | 0.39 [0.24, 0.54] |
|  |  | Outer | 3.09 [2.43, 3.67] | 0.10 [0.08, 0.12] | 0.32 [0.16, 0.48] |
| V2 | Temporal | Inner | 4.11 [3.29, 4.79] | 0.11 [0.09, 0.14] | 0.82 [0.74, 0.88] |
|  |  | Outer | 1.34 [1.11, 1.54] | 0.02 [0.02, 0.03] | 0.61 [0.29, 0.78] |
| V2 | Superior | Inner | 2.08 [1.60, 2.50] | 0.08 [0.06, 0.10] | 0.65 [0.53, 0.76] |
|  |  | Outer | 2.17 [1.61, 2.66] | 0.06 [0.05, 0.08] | 0.44 [0.24, 0.61] |
| V2 | Inferior | Inner | 2.70 [2.07, 3.30] | 0.12 [0.09, 0.15] | 0.43 [0.28, 0.57] |
|  |  | Outer | 1.92 [1.51, 2.29] | 0.06 [0.05, 0.07] | 0.46 [0.21, 0.65] |

### **Supplementary Table S4. Inter-Visit Reliability of Inner- and Outer Peau d’Orange Boundary**

| **Meridian** | **Boundary** | **Standard Error of Measurement** | **Coefficient of Variation** | **Intraclass Correlation Coefficient (two-way random, agreement)** |
| --- | --- | --- | --- | --- |
| Temporal | Inner | 1.97 [1.57, 2.29] | 0.05 [0.04, 0.06] | 0.95 [0.93, 0.97] |
|  | Outer | 1.12 [0.85, 1.36] | 0.02 [0.02, 0.02] | 0.86 [0.78, 0.91] |
| Superior | Inner | 1.28 [0.99, 1.52] | 0.05 [0.04, 0.06] | 0.86 [0.79, 0.91] |
|  | Outer | 1.74 [1.24, 2.23] | 0.05 [0.04, 0.06] | 0.63 [0.47, 0.75] |
| Inferior | Inner | 1.46 [1.12, 1.76] | 0.07 [0.05, 0.08] | 0.78 [0.67, 0.86] |
|  | Outer | 1.69 [1.24, 2.09] | 0.05 [0.04, 0.06] | 0.68 [0.53, 0.79] |

### **Supplementary Table S5. Inter-Eye Concordance of Inner- and Outer Peau d’Orange Boundary**

| **Meridian** | **Boundary** | **Standard Error of Measurement** | **Coefficient of Variation** | **Intraclass Correlation Coefficient (two-way random, agreement)** |
| --- | --- | --- | --- | --- |
| Temporal | Inner | 3.52 [2.89, 4.06] | 0.09 [0.08, 0.11] | 0.85 [0.77, 0.90] |
|  | Outer | 1.54 [1.12, 1.96] | 0.03 [0.02, 0.04] | 0.73 [0.60, 0.82] |
| Superior | Inner | 1.56 [1.20, 1.86] | 0.06 [0.05, 0.07] | 0.81 [0.70, 0.88] |
|  | Outer | 1.96 [1.52, 2.34] | 0.06 [0.04, 0.07] | 0.50 [0.31, 0.66] |
| Inferior | Inner | 2.36 [1.86, 2.79] | 0.11 [0.08, 0.13] | 0.46 [0.25, 0.62] |
|  | Outer | 2.47 [1.98, 2.88] | 0.08 [0.06, 0.09] | 0.32 [0.10, 0.51] |

### **Supplementary Table S6. Linear Regression Models for Peau d’Orange Boundary Extend in Association with Age by Retinal Sector.** For each model, the table lists the estimated intercept and age effect (Estimate, 95% CI) with corresponding *p*-values.

| **Peau d’orange Boundary** | **Predictors** | **Estimates** | **95% CI** | **p** |
| --- | --- | --- | --- | --- |
| Temporal inner | (Intercept) | 15.17 | 2.01 – 28.32 | **0.024** |
|  | Age [deg/decade] | 4.35 | 1.81 – 6.89 | **0.001** |
| Temporal outer | (Intercept) | 56.16 | 51.10 – 61.22 | **<0.001** |
|  | Age [deg/decade] | -0.28 | -1.19 – 0.64 | 0.556 |
| Superior inner | (Intercept) | 20.34 | 14.94 – 25.74 | **<0.001** |
|  | Age [deg/decade] | 1 | -0.04 – 2.05 | 0.06 |
| Superior outer | (Intercept) | 36.05 | 31.48 – 40.62 | **<0.001** |
|  | Age [deg/decade] | -0.11 | -0.94 – 0.72 | 0.788 |
| Inferior inner | (Intercept) | 18.26 | 13.67 – 22.84 | **<0.001** |
|  | Age [deg/decade] | 0.79 | -0.07 – 1.65 | 0.071 |
| Inferior outer | (Intercept) | 34.43 | 30.09 – 38.77 | **<0.001** |
|  | Age [deg/decade] | -0.43 | -1.17 – 0.32 | 0.259 |

### **Supplementary Table S7. Mixed-effects models for Rod Intercept Time (RIT) delay in Association with Peau d’Orange Temporal Inner Boundary.** Linear mixed-effects models were fitted for each eccentricity (8°, 15°, 30°, 46°) and evaluated based on the temporal inner boundary extend and age. PID was included as a random effect.

| **Temporal Inner Boundary** | **RIT at 8°** | | | **RIT at 15°** | | | **RIT at 30°** | | | **RIT at 46°** | | |
| --- | --- | --- | --- | --- | --- | --- | --- | --- | --- | --- | --- | --- |
| **Predictors** | **Estimates** | **95% CI** | **p** | **Estimates** | **95% CI** | **p** | **Estimates** | **95% CI** | **p** | **Estimates** | **95% CI** | **p** |
| (Intercept) | -24.49 | -50.98 – 2.00 | 0.069 | -16.84 | -38.75 – 5.07 | 0.129 | -3.93 | -16.87 – 9.00 | 0.544 | 6.31 | -6.57 – 19.19 | 0.329 |
| Extent [min/degree] | 1.05 | 0.41 – 1.70 | **0.002** | 0.91 | 0.37 – 1.44 | **0.001** | 0.14 | -0.22 – 0.50 | 0.435 | 0.10 | -0.26 – 0.46 | 0.591 |
| Age [min/decade] | 3.18 | -2.37 – 8.73 | 0.254 | 0.83 | -3.76 – 5.41 | 0.719 | 2.60 | -0.17 – 5.36 | 0.065 | 0.84 | -1.92 – 3.60 | 0.545 |
| **Random Effects** | | | | | | | | | | | | |
| σ^2^ | 33.66 | | | 23.14 | | | 43.17 | | | 50.97 | | |
| τ_00_ | 172.31 _PID_ | | | 117.74 _PID_ | | | 21.53 _PI_D | | | 17.03 _PID_ | | |
| ICC | 0.84 | | | 0.84 | | | 0.33 | | | 0.25 | | |
| N | 26 _PID_ | | | 26 _PID_ | | | 26 _PID_ | | | 26 _PID_ | | |
| Observations | 52 | | | 52 | | | 52 | | | 51 | | |
| Marginal R^2^ / Conditional R^2^ | 0.401 / 0.902 | | | 0.341 / 0.892 | | | 0.184 / 0.455 | | | 0.037 / 0.278 | | |
| σ^2^ = residual variance, τ_00_ = between subject variance, ICC = intraclass correlation coefficient, N = number of participants | | | | | | | | | | | | |

### **Supplementary Table S8. Mixed-effects models for Rod Intercept Time (RIT) delay in Association with Peau d’Orange Superior Inner Boundary.** Linear mixed-effects models were fitted for each eccentricity (8°, 15°, 30°, 46°) and evaluated based on the superior inner boundary extend and age. PID was included as a random effect.

| **Superior Inner Boundary** | **RIT at 8°** | | | **RIT at 15°** | | | **RIT at 30°** | | | **RIT at 46°** | | |
| --- | --- | --- | --- | --- | --- | --- | --- | --- | --- | --- | --- | --- |
| **Predictors** | **Estimates** | **95% CI** | **p** | **Estimates** | **95% CI** | **p** | **Estimates** | **95% CI** | **p** | **Estimates** | **95% CI** | **p** |
| (Intercept) | -31.99 | -71.07 – 7.10 | 0.106 | -26.91 | -58.58 – 4.77 | 0.094 | -17.61 | -37.34 – 2.13 | 0.079 | 0.32 | -20.28 – 20.92 | 0.975 |
| Extent [min/degree] | 1.16 | -0.05 – 2.38 | 0.059 | 1.18 | 0.13 – 2.23 | **0.028** | 0.79 | -0.02 – 1.60 | 0.055 | 0.37 | -0.47 – 1.22 | 0.381 |
| Age [min/decade] | 6.56 | 0.47 – 12.64 | **0.035** | 3.53 | -1.20 – 8.26 | 0.140 | 2.36 | -0.02 – 4.74 | 0.052 | 0.86 | -1.60 – 3.32 | 0.486 |
| **Random Effects** | | | | | | | | | | | | |
| σ^2^ | 23.97 | | | 19.76 | | | 40.38 | | | 49.27 | | |
| τ_00_ | 280.27 _PID_ | | | 164.82 _PID_ | | | 20.39 _PID_ | | | 18.57 _PID_ | | |
| ICC | 0.92 | | | 0.89 | | | 0.34 | | | 0.27 | | |
| N | 26 _PID_ | | | 26 _PID_ | | | 26 _PID_ | | | 26 _PID_ | | |
| Observations | 52 | | | 52 | | | 52 | | | 51 | | |
| Marginal R^2^ / Conditional R^2^ | 0.233 / 0.940 | | | 0.188 / 0.913 | | | 0.239 / 0.494 | | | 0.048 / 0.308 | | |
| σ^2^ = residual variance, τ_00_ = between subject variance, ICC = intraclass correlation coefficient, N = number of participants | | | | | | | | | | | | |

### **Supplementary Table S9. Mixed-effects models for Rod Intercept Time (RIT) delay in Association with Peau d’Orange Inferior Inner Boundary.** Linear mixed-effects models were fitted for each eccentricity (8°, 15°, 30°, 46°) and evaluated based on the inferior inner boundary extend and age. PID was included as a random effect.

| **Inferior Inner Boundary** | **RIT at 8°** | | | **RIT at 15°** | | | **RIT at 30°** | | | **RIT at 46°** | | |
| --- | --- | --- | --- | --- | --- | --- | --- | --- | --- | --- | --- | --- |
| **Predictors** | **Estimates** | **95% CI** | **p** | **Estimates** | **95% CI** | **p** | **Estimates** | **95% CI** | **p** | **Estimates** | **95% CI** | **p** |
| (Intercept) | -4.14 | -43.11 – 34.83 | 0.832 | -6.74 | -38.68 – 25.21 | 0.673 | -14.15 | -34.31 – 6.01 | 0.164 | 10.11 | -10.35 – 30.56 | 0.325 |
| Extent [min/degree] | -0.25 | -1.35 – 0.86 | 0.657 | 0.18 | -0.80 – 1.16 | 0.714 | 0.65 | -0.21 – 1.51 | 0.135 | -0.13 | -1.01 – 0.76 | 0.777 |
| Age [min/decade] | 7.98 | 1.57 – 14.40 | **0.016** | 4.68 | -0.40 – 9.76 | 0.070 | 2.75 | 0.38 – 5.13 | **0.024** | 1.35 | -1.01 – 3.71 | 0.256 |
| **Random Effects** | | | | | | | | | | | | |
| σ^2^ | 24.03 | | | 19.84 | | | 40.07 | | | 51.10 | | |
| τ_00_ | 323.46 _PID_ | | | 199.69 _PID_ | | | 23.70 _PID_ | | | 17.23 _PID_ | | |
| ICC | 0.93 | | | 0.91 | | | 0.37 | | | 0.25 | | |
| N | 26 _PID_ | | | 26 _PID_ | | | 26 _PID_ | | | 26 _PID_ | | |
| Observations | 52 | | | 52 | | | 52 | | | 51 | | |
| Marginal R^2^ / Conditional R^2^ | 0.188 / 0.944 | | | 0.122 / 0.921 | | | 0.209 / 0.503 | | | 0.032 / 0.276 | | |
| σ^2^ = residual variance, τ_00_ = between subject variance, ICC = intraclass correlation coefficient, N = number of participants | | | | | | | | | | | | |

### **Supplementary Table S10. Multiple Hypothesis Testing Using the Benjamini–Hochberg (BH) Procedure**

Multiple hypothesis testing was addressed using the Benjamini–Hochberg (BH) procedure to control the false discovery rate (FDR). Specifically, we extracted the raw p-values for the fixed effects of interest (effect of extent) from the mixed-effects models and applied BH correction. We report both the raw p-values and the corresponding FDR-adjusted q-values in Supplementary Tables S5–S7. Covariate effects (e.g., AgeDecades) were included for confounding adjustment but were not considered part of the tested hypothesis family and therefore not subjected to FDR correction. Conclusions were based on q < 0.05 as the threshold for statistical significance.

| **RIT Eccentricity**  **(Outcome)** | **Meridian** | **Boundary** | **Est [95% CI]** | **p raw** | **q BH** | **Significance Bonferroni** | **Significance Benjamini–Hochberg** |
| --- | --- | --- | --- | --- | --- | --- | --- |
| 8 | Temporal | Inner | **1.05 [0.41, 1.70]** | **0.002** | **0.004** |  | ***** |
| 15 | Temporal | Inner | **0.91 [0.37, 1.44]** | **0.001** | **0.004** | **†** | ***** |
| 30 | Temporal | Inner | 0.14 [-0.22, 0.50] | 0.438 | 0.584 |  |  |
| 46 | Temporal | Inner | 0.10 [-0.27, 0.46] | 0.593 | 0.593 |  |  |
| 8 | Superior | Inner | 1.16 [-0.05, 2.38] | 0.06 | 0.081 |  |  |
| 15 | Superior | Inner | 1.18 [0.13, 2.24] | 0.029 | 0.081 |  |  |
| 30 | Superior | Inner | 0.79 [-0.02, 1.60] | 0.057 | 0.081 |  |  |
| 46 | Superior | Inner | 0.37 [-0.48, 1.23] | 0.383 | 0.383 |  |  |
| 8 | Inferior | Inner | -0.25 [-1.36, 0.87] | 0.658 | 0.778 |  |  |
| 15 | Inferior | Inner | 0.18 [-0.81, 1.17] | 0.714 | 0.778 |  |  |
| 30 | Inferior | Inner | 0.65 [-0.21, 1.52] | 0.136 | 0.545 |  |  |
| 46 | Inferior | Inner | -0.13 [-1.02, 0.77] | 0.778 | 0.778 |  |  |

### **Supplementary Table S11. Sensitivity Analyses**

To assess whether the association between Peau d’orange extent and rod-intercept time (RIT) varied with age, we performed a stratified analysis, splitting participants at the median age of 55.4 years (Younger vs. Older). Only for the younger stratum, Peau d’orange extent remained significantly associated with delayed RIT. The effect size was greater in younger patients than in older patients. This difference is most plausibly explained by a methodological limitation of the study: dark adaptation testing was capped at 60 minutes. This ceiling effect likely truncated the measurable range of delay in older patients, thereby attenuating the observed strength of association.

| **RIT Eccentricity** | **Boundary** | **Meridian** | **Age Stratum** | **Estimate [min/deg]** | **p raw** |
| --- | --- | --- | --- | --- | --- |
| **(Outcome)** |  |  |  |  |  |
| 8 | Inner | Temporal | Younger | **1.43 [0.75, 2.11]** | **<0.001** |
| 8 | Inner | Temporal | Older | 0.73 [-0.40, 1.87] | 0.194 |
| 8 | Inner | Superior | Younger | 0.45 [-0.41, 1.31] | 0.274 |
| 8 | Inner | Superior | Older | 1.20 [-1.11, 3.51] | 0.293 |
| 8 | Inner | Inferior | Younger | -0.74 [-1.50, 0.02] | 0.055 |
| 8 | Inner | Inferior | Older | -0.07 [-1.98, 1.85] | 0.944 |
| 15 | Inner | Temporal | Younger | **1.02 [0.42, 1.62]** | **0.003** |
| 15 | Inner | Temporal | Older | 0.21 [-0.37, 0.79] | 0.442 |
| 15 | Inner | Superior | Younger | **1.95 [0.35, 3.55]** | **0.019** |
| 15 | Inner | Superior | Older | 1.00 [-0.12, 2.11] | 0.077 |
| 15 | Inner | Inferior | Younger | -0.01 [-1.88, 1.87] | 0.994 |
| 15 | Inner | Inferior | Older | 0.43 [-0.42, 1.28] | 0.292 |
| 30 | Inner | Temporal | Younger | 0.06 [-0.18, 0.29] | 0.622 |
| 30 | Inner | Temporal | Older | 0.75 [-0.04, 1.54] | 0.062 |
| 30 | Inner | Superior | Younger | 0.50 [-0.04, 1.04] | 0.07 |
| 30 | Inner | Superior | Older | 1.37 [-0.28, 3.02] | 0.097 |
| 30 | Inner | Inferior | Younger | 0.02 [-0.63, 0.66] | 0.961 |
| 30 | Inner | Inferior | Older | 1.46 [-0.09, 3.02] | 0.064 |
| 46 | Inner | Temporal | Younger | -0.02 [-0.30, 0.25] | 0.845 |
| 46 | Inner | Temporal | Older | 0.52 [-0.25, 1.29] | 0.172 |
| 46 | Inner | Superior | Younger | 0.59 [-0.14, 1.33] | 0.106 |
| 46 | Inner | Superior | Older | 0.23 [-1.40, 1.86] | 0.765 |
| 46 | Inner | Inferior | Younger | 0.14 [-0.70, 0.98] | 0.73 |
| 46 | Inner | Inferior | Older | -0.23 [-1.78, 1.32] | 0.761 |

### **Supplementary Table S12. Non-linear Mixed Model with a Step Function to Describe the Relationship Between the Temporal Inner Peau d’Orange Boundary Extent and the Rod-Intercept Time (RIT)**

|  | **RIT at 8°** | | **RIT at 15°** | |
| --- | --- | --- | --- | --- |
| **Predictors** | **Estimates** | **Credible Interval (95%)** | **Estimates** | **Credible Interval (95%)** |
| Intercept [min] | 10.14 | 5.53 – 14.78 | 9.32 | 5.02 – 13.66 |
| Age [min/decade] | 0.79 | 0.36 – 1.72 | 0.73 | 0.34 – 1.49 |
| Effect of extent [min/mm] | 1.48 | 0.76 – 2.34 | 1.27 | 0.50 – 2.75 |
| Critical extent [mm] | 27.15 | 17.06 – 33.09 | 32.56 | 22.64 – 40.56 |
| Observations | 52 | | 52 | |
| Marginal R2 / Conditional R2 | 0.440 / 0.902 | | 0.328 / 0.891 | |

### **Supplementary Methods. Non-linear Mixed Model with a Step Function**

To describe the non-linear relationship between the temporal inner Peau d’orange boundary extent and the rod-intercept time at 8° and 15° eccentricity, we fitted a Bayesian non-linear mixed-model (estimated using MCMC sampling with four chains of 2000 iterations and a warmup of 1000).

Formula: RIT ~ b0 + b1 × Age + b2 × (Extent - Omega) × step (Extent - Omega)

The following priors were used:

| **Parameter** | **Description** | **Prior** | **95% HDI of the Prior** | **Reasoning** |
| --- | --- | --- | --- | --- |
| b0 | Intercept | normal (10, 2.5) | [5, 15] | Weakly informative prior based on RIT normal data |
| b1 | Effect of Age | lognormal (-0.3, 0.4) | [0.27, 1.47] | Weakly informative prior based on ageing effect on RIT (per decade)^4,5^ |
| b2 | Effect of Peau d’Orange Extent | normal (0, 1.5) | [-3, 3] | Uninformative prior |
| Omega | Critical Extent/Step Function | normal (35, 10) | [15, 55] | Weakly informative prior based on the age range of patients in the study |
